# Psychotropic Receptor Modulation and Cognition in Psychotic Disorders

**DOI:** 10.64898/2026.10.12.26365151

**Authors:** Cameron J Watson, Jeremy Hall, Richard SE Keefe, Philip McGuire, Sridhar Natesan, Maxime Taquet, Rachel Upthegrove, James TR Walters, Toby Pillinger, Robert A McCutcheon

## Abstract

**Background:** Cognitive impairment is a major determinant of functional outcome in psychotic disorders, yet the contribution of psychotropic medication is incompletely understood. This study aimed to determine whether personalised receptor-engagement profiles derived from prescribed psychotropic medications are associated with cognitive performance in individuals with psychotic disorders.

**Methods:** We performed a cross-sectional study using clinical, cognitive, and medication data from the Bipolar-Schizophrenia Network on Intermediate Phenotypes studies. In 1,067 (51.5% female) individuals with psychotic disorders, we estimated personalised receptor-engagement profiles based on the prescribed dose and known pharmacological properties of 49 psychotropic medications across 32 drug targets. Correlated receptor-target modulation profiles were summarized into seven pharmacological clusters. Extreme gradient boosting models were trained using repeated nested cross-validation, with the contribution of pharmacological profiles to predicted cognition quantified using out-of-sample SHapley Additive exPlanations (SHAP) values. Overall pharmacological contribution was tested using within-diagnosis permutation testing.

**Results:** Estimated receptor-engagement profiles were significantly associated with cognitive performance independent of diagnosis. A receptor profile involving alpha2-adrenergic, dopaminergic (DRD4, DRD5) and several serotonergic receptors was associated with poorer cognitive performance, whereas dopamine transporter (SLC6A3) engagement was associated with better cognitive performance. Muscarinic receptor modulation was additionally associated with poorer verbal memory and motor speed, while the DRD2-containing receptor profile was not associated with cognitive performance after correction for multiple testing.

**Conclusions:** Modelling psychotropic medications according to receptor pharmacology identified receptor-engagement profiles associated with cognitive performance in psychotic disorders. This framework may help identify receptor systems for future investigation as potential targets for cognition-enhancing therapies and support the development of personalised prescribing strategies.

## Introduction

Cognitive impairment is a core feature of psychotic disorders and is strongly associated with poor functional outcomes^1^. In schizophrenia, impairments span multiple cognitive domains and, when combined globally, approach 1.5 SDs below the performance of healthy controls^1^. Cognitive impairment is also increasingly recognized in bipolar disorder with psychosis, supporting the view that cognitive dysfunction represents a transdiagnostic feature of psychotic illness^2^. Crucially, cognition is not uniformly impaired across individuals with psychotic disorders, particularly at first episode, where patients show greater variability in cognitive performance than controls^3^. This heterogeneity highlights the need for more individualized approaches to understanding cognitive dysfunction.

Cognitive performance in psychotic disorders reflects multiple influences, including genetic liability, environmental exposures, illness characteristics, and antipsychotic treatment^2,4,5^. Cognitive side effects are associated with several antipsychotics and are an important driver of both treatment-related distress and a desire to discontinue medication^6^. Whilst antipsychotics primarily act via dopaminergic mechanisms, they differ markedly in their affinities across other receptor systems. Furthermore, many individuals with psychotic disorders are prescribed combinations of other psychotropic medications including antidepressants, mood stabilizers, and hypnotics, which may also influence cognition through distinct pharmacological mechanisms^7–9^. Currently, treatment selection rarely incorporates individual neuropharmacological profiles or considers how specific receptor engagement may relate to cognitive outcomes.

In this study, we estimated the extent to which each participant’s prescribed psychotropic medications engaged 32 neurotransmitter receptors based on medication dose and known receptor pharmacology. We then used machine-learning to examine whether these receptor-engagement profiles were associated with cognition. Finally, we identified the receptor profiles most strongly associated with cognitive performance across domains. This approach was designed to test whether receptor-informed pharmacological profiles contribute to cognitive heterogeneity in individuals with psychotic disorders and to support future development of more personalised treatment strategies.

## Methods and Materials

### Participants

Clinical, cognitive, and medication data were obtained from participants enrolled in the Bipolar-Schizophrenia Network on Intermediate Phenotypes studies (BSNIP-1^10^ and BSNIP-2^11^). Participants were recruited between 2008-2020 across six sites in the United States. All participants provided written informed consent, with institutional review board approval obtained at participating sites. BSNIP cohorts included individuals with psychotic disorders (schizophrenia, schizoaffective disorder, or bipolar I disorder with psychosis) as well as healthy controls. Participants with psychosis were included in the current study if they had available cognitive data, medication records, and sufficient pharmacological information to estimate receptor-level modulation.

### Cognitive Assessment

Cognition was assessed using the Brief Assessment of Cognition in Schizophrenia (BACS)^12^, covering six cognitive domains: verbal memory, working memory, motor speed, verbal fluency, attention and processing speed, and executive function. Domain-specific z-scores were calculated using the healthy control group mean and standard deviation as the reference. A composite cognitive score was calculated as the mean of the domain-specific z-scores, dividing by the SD of the control mean z-scores. Participants with composite cognitive scores >5 standard deviations from the mean were excluded, while the most extreme 0.5% of the remaining composite scores were winsorized, with values capped at the corresponding 0.5^th^ and 99.5^th^ percentile boundaries.

### Medication Data

Medication records were harmonised across BSNIP-1 (N=543) and BSNIP-2 (N=524), yielding a cohort of 1,067 participants. In BSNIP-1, medications were retained if recorded as ongoing at assessment. In BSNIP-2, medications were additionally required to be recorded as taken at the time of cognitive testing, with this information unavailable in BSNIP-1. Psychotropics were considered for modelling if taken daily, with likely depot/long-acting injectable preparations excluded. Medication names were standardised and duplicate participant-drug records resolved to estimate total daily dose. To limit the influence of likely duplicate or implausible entries, summed doses above a prespecified drug-specific maximum were capped at that maximum, while single recorded doses were retained up to twice the maximum and capped thereafter.

### Pharmacological Modelling

We modelled receptor engagement across commonly prescribed psychotropic medications (Figure 1), including antipsychotics, antidepressants and stimulants (Supplementary Table 1). Drug-target binding affinity data were obtained primarily from the Psychoactive Drug Screening Program (PDSP) database. Where PDSP provided only a non-specific parent-family affinity for a drug, subtype-specific affinities were supplemented from the literature. Non-specific parent receptor labels (e.g., ADRA1, ADRA2, HTR2 and CHRM) were then excluded from the final target matrix, and receptor labels were harmonized to gene-level nomenclature where appropriate. For each drug-target pair, the median affinity value was used when multiple estimates were available, and interactions with Ki ≥1000 nM were treated as unavailable. Receptors were retained only where usable pharmacological data were available for at least five psychotropics, reducing the likelihood that receptor-level effects reflected individual drugs rather than shared target engagement. The final FMI matrix comprised 49 drugs and 32 receptor targets.

**Figure 1.**
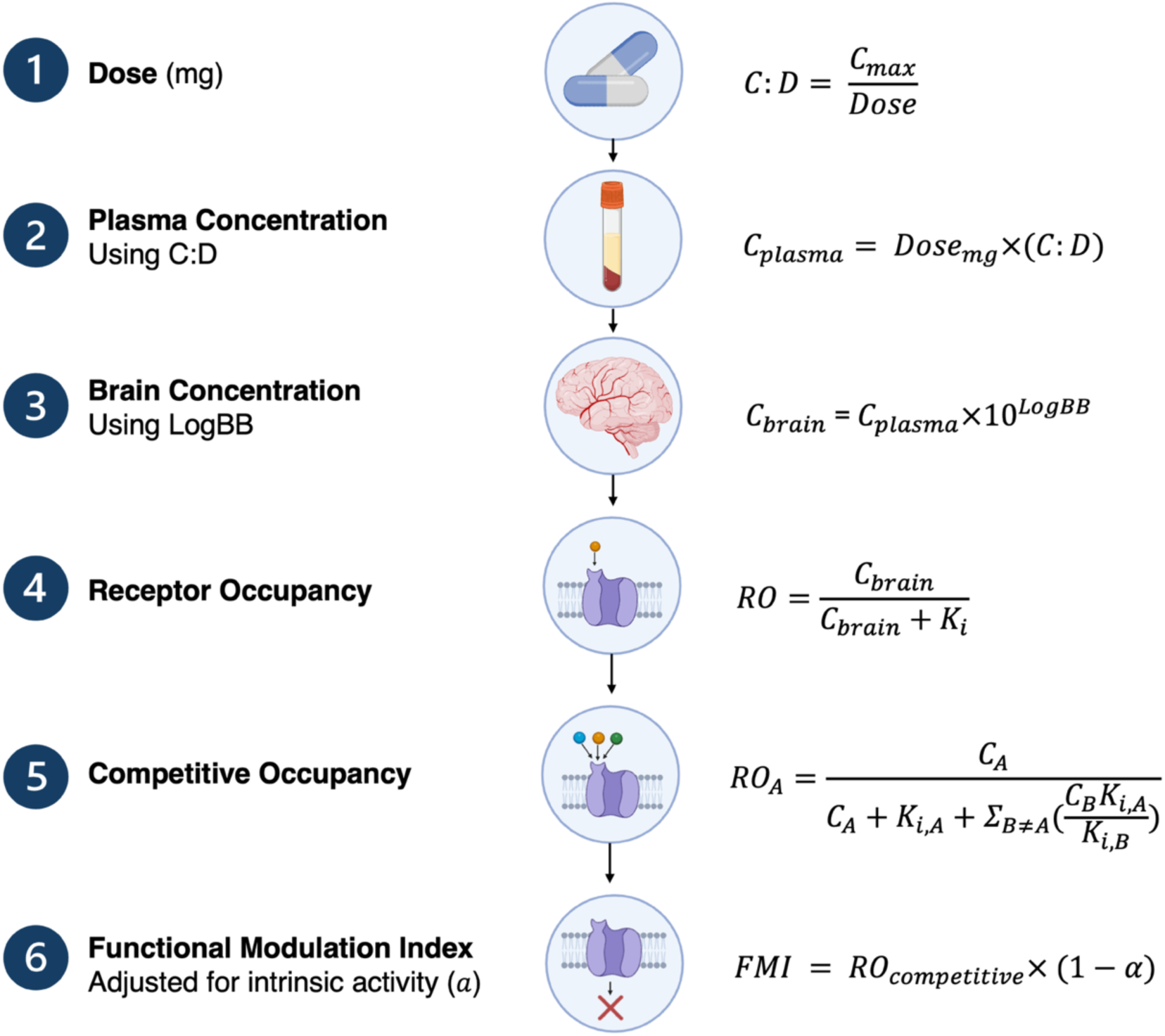
Pipeline for estimating participant-level receptor modulation profiles. Recorded daily medication dose was converted to estimated plasma concentration using drug-specific peak plasma concentration-to-dose (C:D) coefficients. Plasma concentrations were converted to molar units using molecular weight and adjusted for predicted blood-brain barrier penetration (LogBB) to estimate brain concentrations. Drug-target receptor occupancy was estimated from brain concentration and binding affinity (*K_i_*); for participants prescribed multiple drugs acting at the same target, competitive occupancy accounted for simultaneous receptor competition. Competitive occupancy was weighted by intrinsic activity (*a*) to derive the Functional Modulation Index (FMI), with antagonists/inhibitors contributing maximally and full agonists contributing no antagonist-like FMI. Created in BioRender. Watson, C. (2026) https://BioRender.com/6xsadwt

Participant-level medication dose was converted to estimated plasma concentration using drug-specific dose-plasma coefficients using the following steps. Drug-specific peak plasma concentrations (*C_max_*) were obtained from the NIH Drug Insights database and dose-plasma concentration ratio was then calculated as:

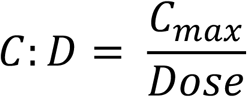

Assuming linear pharmacokinetics, for each prescribed medication, the estimated plasma concentration was calculated from the participant’s recorded dose and the drug-specific C:D ratio.

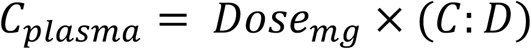

Plasma concentrations were converted to molar units using molecular weight (obtained from PubChem), adjusted for estimated blood-brain barrier penetration (*logBB* values derived from molecular SMILES strings via LogBBpred^13^), with brain concentrations in nmol/L estimated as:

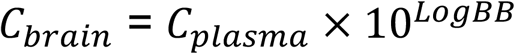

### Receptor Occupancy Modelling

For each participant, drug, and receptor, receptor occupancy was estimated using the drug’s estimated brain concentration and receptor binding affinity using the classical pharmacological receptor-occupancy relationship^14^:

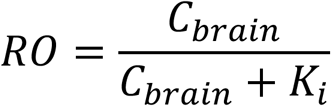

where *C_brain_*is the estimated brain concentration and *K_i_*is the drug-receptor binding affinity, both expressed in nmol/L.

For participants prescribed multiple drugs acting at the same receptor, a competitive binding model was applied to account for simultaneous receptor competition^15^:

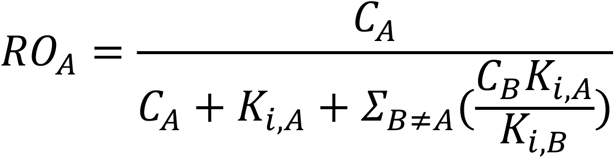

where *C_A_* and *K_i_*_,*A*_ are the estimated brain concentration and affinity of drug *A*, and the summation term represents the competitive contribution of other prescribed drugs (e.g. drug *B*) acting at the same receptor. This produces a competitive occupancy estimate for each participant-drug-receptor combination.

### Functional Modulation Index

To account for the functional direction of drug-target interactions, competitive occupancy estimates were weighted using an intrinsic activity coefficient (*α*). Literature-derived *α* values >0 were applied where partial or full agonist activity was identified (Supplementary Table 3), otherwise the value was set to 0. Values ranged from 0, representing maximal antagonist/inhibitor-like contribution, to 1, representing full agonism.

Functional Modulation Index (FMI) was defined as:

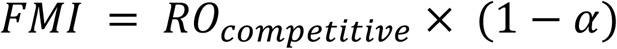

Under this definition, antagonists contribute maximally to FMI, partial agonists contribute proportionally less, and full agonists (*α* = 1) do not contribute. For each participant and receptor, FMI values were summed across all retained medications:

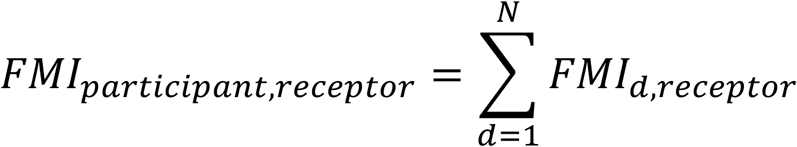

Summed receptor-level FMI values were truncated at 1.0 to prevent values exceeding complete receptor modulation.

### Pharmacological clustering

As receptor-level FMI features were highly correlated, receptors were grouped according to the similarity of their participant-level modulation profiles. Among medicated participants with at least one non-zero FMI value, pairwise Pearson correlations between receptor-level FMI vectors were converted to dissimilarities using 1 − *r*, and average-linkage hierarchical clustering applied to the resulting dissimilarity matrix. The dendrogram was cut at a height of 0.60, and the resulting receptor-to-cluster mapping applied to the full dataset. Participant-level cluster FMI scores were calculated as the mean FMI across constituent receptors, with zero assigned where no receptor-level pharmacological signal was present.

### Predictive modelling of cognitive performance

Predictive modelling was performed using gradient-boosted decision trees implemented with XGBoost^16^. The primary outcome was composite cognitive performance, with secondary analyses of the six BACS domains. Pharmacological predictors were cluster-level FMI indices. Models additionally included demographic, parental, childhood-trauma, substance-use, education, and WRAT reading predictors as previously used in McCutcheon et al^2^ (Supplementary Table 6). Given that substance use, education, and reading abilities could reasonably be a consequence as well as an antecedent of a psychotic disorder, analyses were repeated with these variables excluded. An additional model included PANSS positive, negative, and general psychopathology scores to account for current symptom severity. Numeric predictors were mean-imputed and standardized; categorical predictors were mode-imputed and one-hot encoded. All preprocessing was performed within cross-validation training folds. Hyperparameters were tuned using nested cross-validation. The modal optimal parameter set was then applied in 10-fold cross-validation repeated 10 times, with preprocessing and model fitting restricted to training folds and SHAP values calculated in held-out participants. Participant-level SHAP values were averaged across repeats, and feature importance summarised using mean absolute SHAP values for individual predictors and predictor domains.

To test whether pharmacological features carried individual-level cognition-relevant information, the complete cluster-level FMI profile was permuted as a block across participants within diagnostic group. This preserved diagnosis-level medication structure while breaking the participant-level relationship between receptor modulation and cognition. For each of 500 permutations, the model was refitted and out-of-sample SHAP attribution was recalculated for each pharmacological cluster. Total pharmacological attribution was calculated as the summed mean absolute SHAP attribution across clusters. observed total and cluster-specific attribution values were compared with their corresponding permutation-derived null distributions using one-sided empirical p-values. Cluster-specific p-values were corrected across the seven pharmacological clusters using the Benjamini-Hochberg false discovery rate.

## Results

### Cohort Characteristics

The analytic patient sample included 1,067 participants: 432 with schizophrenia, 353 with schizoaffective disorder, and 282 with bipolar disorder with psychosis (Table 1).

**Table 1.** Clinical and Demographic Characteristics of the Study Sample. : Educational variables were coded using the Hollingshead educational scale; higher scores indicate lower educational attainment. WRAT denotes Wide Range Achievement Test 4 reading score. CTQ denotes Childhood Trauma Questionnaire. Substance-use variables reflect lifetime reported use. Antipsychotic polypharmacy refers to more than one prescribed antipsychotic at assessment. Percentages are calculated among participants with available data for the relevant variable.

| Characteristic | Overall | Schizophrenia | Schizoaffective disorder | Bipolar with psychosis |
| --- | --- | --- | --- | --- |
| No. participants | 1,067 | 432 | 353 | 282 |
| Age, mean (SD), years | 37.90 (12.37) | 38.33 (12.69) | 38.99 (11.96) | 35.88 (12.18) |
| Female sex, No. (%) | 549 (51.5%) | 159 (36.8%) | 208 (58.9%) | 182 (64.5%) |
| White, No. (%) | 512 (48.0%) | 169 (39.1%) | 150 (42.5%) | 193 (68.4%) |
| Black or African American, No. (%) | 375 (35.1%) | 192 (44.4%) | 134 (38.0%) | 49 (17.4%) |
| Hispanic, No. (%) | 111 (10.4%) | 40 (9.3%) | 47 (13.3%) | 24 (8.5%) |
| Other race, No. (%) | 69 (6.5%) | 31 (7.2%) | 22 (6.2%) | 16 (5.7%) |
| Composite cognition score, mean (SD) | -1.17 (1.16) | -1.43 (1.14) | -1.24 (1.11) | -0.69 (1.11) |
| Participant education scale, mean (SD) | 3.30 (1.14) | 3.49 (1.15) | 3.42 (1.13) | 2.86 (1.04) |
| WRAT/reading score, mean (SD) | 57.88 (8.42) | 56.55 (9.06) | 57.13 (8.34) | 60.87 (6.58) |
| Father education scale, mean (SD) | 3.31 (1.70) | 3.37 (1.69) | 3.52 (1.74) | 3.00 (1.61) |
| Mother education scale, mean (SD) | 3.34 (1.54) | 3.40 (1.56) | 3.58 (1.57) | 2.98 (1.42) |
| Maternal age, mean (SD) | 26.73 (6.74) | 26.49 (6.86) | 25.71 (6.50) | 28.93 (6.47) |
| Paternal age, mean (SD) | 30.60 (8.17) | 31.11 (8.51) | 29.49 (8.11) | 31.52 (7.56) |
| CTQ total score, mean (SD) | 52.12 (20.74) | 51.00 (19.62) | 54.96 (21.99) | 48.75 (19.87) |
| Cannabis use ever, No. (%) | 397 (75.8%) | 153 (72.2%) | 158 (77.1%) | 86 (80.4%) |
| Any non-cannabis substance use, No. (%) | 382 (72.9%) | 137 (64.6%) | 155 (75.6%) | 90 (84.1%) |
| Antipsychotic polypharmacy, No. (%) | 139 (13.0%) | 68 (15.7%) | 45 (12.7%) | 26 (9.2%) |

### Pharmacology Features

Psychotropic exposure was transformed into receptor-level Functional Modulation Index (FMI) values across 49 drugs (Supplementary Table 1). After receptor-level coverage filtering, the analytic FMI matrix included 32 receptor and transporter targets. Hierarchical clustering of receptor-receptor FMI correlations identified seven pharmacological clusters (Figure 2; Supplementary Table 4).

**Figure 2.**
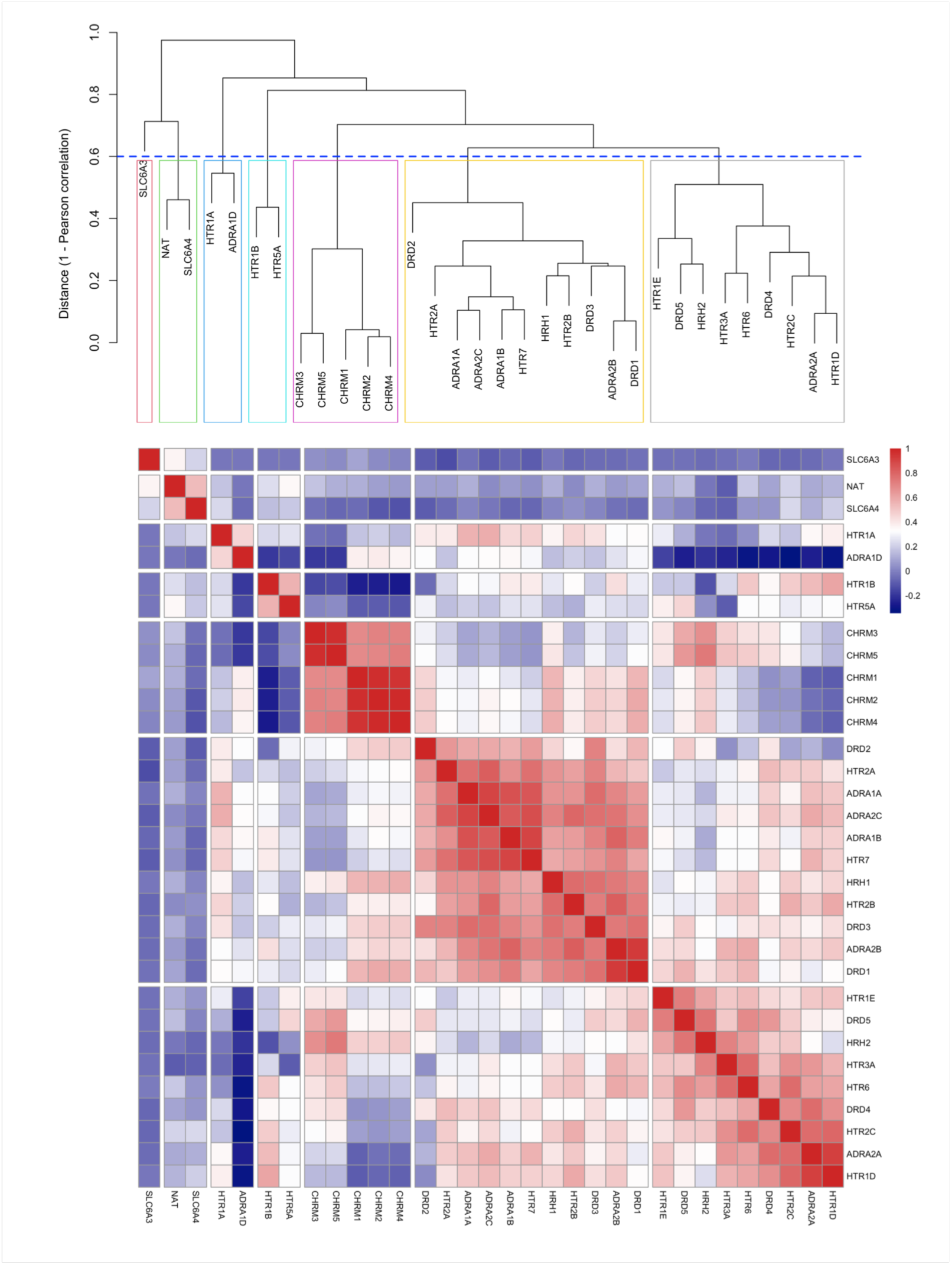
Pharmacological cluster construction from receptor-level FMI profiles. Hierarchical clustering was performed using dissimilarity defined as 1 − *r*, where *r* is the Pearson correlation between participant-level receptor FMI profiles. The dendrogram was cut at a height of 0.60 (blue dashed line), yielding seven pharmacological clusters, indicated by coloured boxes. The heatmap shows pairwise Pearson correlations between receptor-level FMI profiles, with red indicating positive and blue indicating negative correlations.

### Participant FMI Profiles are Associated with Cognition

To assess whether medication-derived receptor modulation carried participant-level cognition-relevant information, FMI cluster profiles were block-permuted across participants within diagnostic groups and models refitted. In the full covariate model, observed total pharmacological attribution was 0.224, compared with a permutation null mean of 0.112 (SD=0.019; range, 0.068– 0.192), a 49.9% reduction overall (p=0.002). The effect was similarly observed in the restricted model excluding education, reading ability, and substance-use variables (observed=0.306; null mean=0.155 [SD=0.023; range, 0.094–0.249]; 49.3% reduction; p=0.002). Model performance metrics are provided in Supplementary Table 10.

### Pharmacological Attribution Localizes to Specific Receptor-Modulation Profiles

Participant-level SHAP distributions revealed that FMI cluster contributions to predicted cognition were heterogeneous in both magnitude and direction (Figure 3A). The clearest negative pattern emerged for Cluster 2, comprising ADRA2A, DRD4, DRD5, HRH2, HTR1D, HTR1E, HTR2C, HTR3A, and HTR6. Higher FMI values in this cluster were consistently associated with negative SHAP contributions, indicating lower predicted composite cognition. Modulation of Clusters 4 (HTR1B and HTR5A) and Cluster 5 (CHRM1-CHRM5) also contributed negatively to cognition. A contrasting pattern was observed for Cluster 7, corresponding to SLC6A3/DAT. This cluster showed a sparse but marked positive SHAP pattern, with higher FMI values associated with higher predicted composite cognition amongst participants with non-zero SLC6A3 modulation.

**Figure 3.**
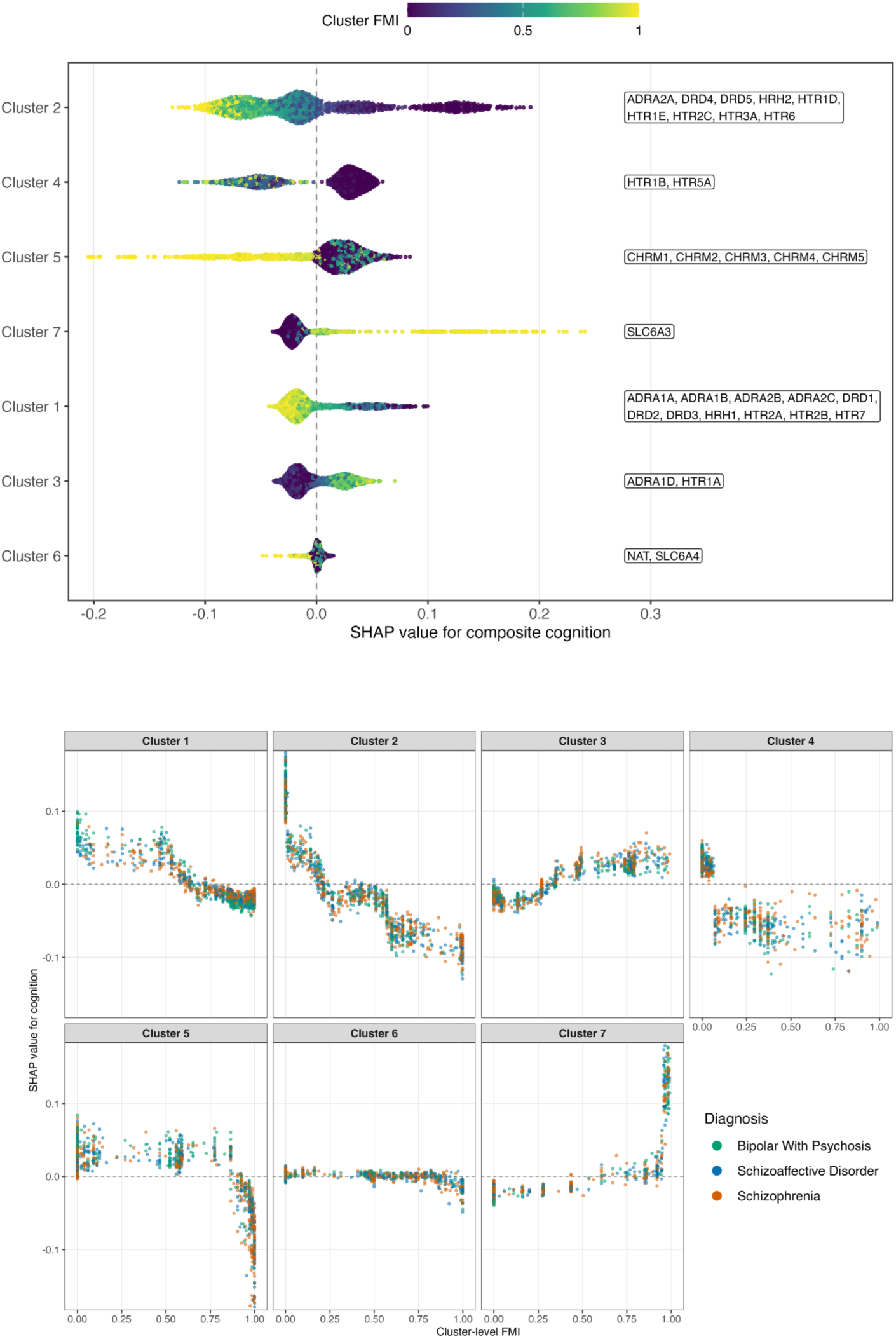
Cluster-level pharmacological attribution for composite cognition. (A) Distribution of participant-level SHAP contributions across FMI clusters, coloured by cluster FMI. (B) Relationship between cluster FMI and SHAP contribution stratified by diagnosis. Each point represents an individual participant. Positive SHAP values indicate contributions toward higher predicted composite cognition, whereas negative values indicate contributions toward lower predicted composite cognition.

We then tested whether cluster-level attribution patterns exceeded a within-diagnosis FMI-shuffle null. In the full model, Cluster 2 showed the strongest cluster-specific attribution, with an observed mean absolute SHAP of 0.059 compared with a permutation null mean of 0.017 (p=0.002; FDRp=0.014; Supplementary Table 7). Cluster 4 also survived correction (observed SHAP 0.041; null mean 0.014; p=0.012; FDR p=0.042), as did Cluster 7, corresponding to SLC6A3, which showed observed attribution of 0.034 compared with a null mean of 0.012 (p=0.020; FDRp=0.047). In the restricted model, Clusters 2 (p=0.006; FDRp=0.021) and 7 (p=0.004; FDRp=0.021) remained significant, whereas Cluster 4 did not survive correction (p=0.034; FDR p=0.073; Supplementary Table 7).

To assess whether FMI-SHAP relationships were explained by symptom severity, we repeated the full model including PANSS positive, negative, and general psychopathology scores as additional predictors. Pharmacological attribution remained significant after permutation (SHAP=0.216; null mean=0.099; 54.0% reduction; p=0.002). Clusters 2 (p=0.002; FDR p=0.014), 4 (p=0.012; FDR p=0.033), and 7 (p=0.014; FDR p=0.033) remained significant after FDR correction (Supplementary Table 7). Diagnosis-stratified FMI-SHAP plots showed similar patterns across psychotic disorders (Figure 3B). This suggests that the main pharmacological attribution patterns were not driven by diagnostic group or illness severity.

### Pharmacological Attribution Varies by Cognitive Domain

Total pharmacological attribution was significantly greater than the permutation null for verbal memory (45.2% reduction; p=0.002), working memory (27.7%; p=0.024), motor speed (36.4%; p=0.004), verbal fluency (53.6%; p=0.002), attention and processing speed (47.5%; p=0.002), but not executive function (11.3%; p=0.222; Figure 4, Supplementary Table 8).

**Figure 4.**
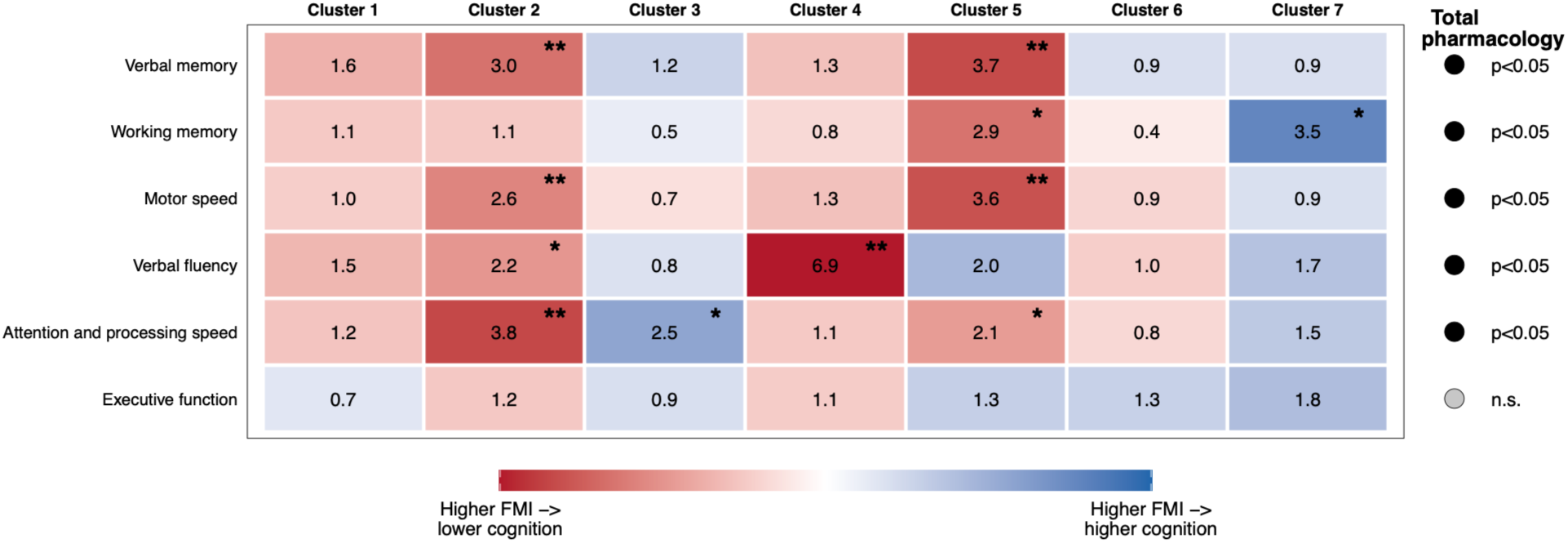
Domain-specific pharmacological attribution across FMI clusters. Tile numbers show the observed mean absolute SHAP attribution divided by the permutation null mean. Darker tiles indicate stronger attribution enrichment. Red indicates that higher FMI values contributed to lower predicted cognition, while blue indicates that higher FMI values contributed to higher predicted cognition. *Uncorrected p<0.05; **FDR-adjusted p<0.05. The right-hand symbols indicate whether total pharmacological attribution across all clusters was significant for each cognitive domain (p<0.05).

Cluster-level analyses showed partially domain-specific patterns. Cluster 2 showed significant negative attribution for verbal memory (p=0.004; FDR p=0.014), motor speed (p=0.014; FDR p=0.049), and attention and processing speed (p=0.002; FDR p=0.014). Cluster 5 showed significant negative attribution for verbal memory (p=0.002; FDR p=0.014) and motor speed (p=0.004; FDR p=0.028), while Cluster 4 showed significant negative attribution for verbal fluency (p=0.002; FDR p=0.014). No individual cluster survived FDR correction for working memory or executive function, although Cluster 5 and Cluster 7 showed suggestive negative (p=0.012; FDR p=0.056) and positive attribution (p=0.016; FDR p=0.056), respectively, for working memory (Figure 4; Supplementary Table 9).

## Discussion

Predicting the impact of pharmacological treatment on cognition is challenging. Both psychotic disorders and psychotropic medications are biologically diverse, yet studies often pool diagnoses or reduce medications to coarse categories or dose-equivalent measures. Taking a novel approach, we derived personalised Functional Modulation Indices (FMI), quantifying the impact of psychotropic medications across receptor systems in individuals with psychotic disorders. FMI profiles carried cognition-relevant pharmacological signal beyond diagnosis, with pharmacological attribution approximately halving after permutation. Analyses localized negative associations between cognition and Cluster 2, comprising adrenergic, dopaminergic and serotonergic targets, and positive associations with Cluster 7, corresponding to SLC6A3, which encodes the dopamine transporter (DAT). These findings were robust to sensitivity analyses excluding education, reading ability and substance-use variables, and following adjustment for symptom severity. Domain-specific analyses additionally identified negative associations between muscarinic receptor modulation and both verbal memory and motor speed.

These findings extend previous work in BSNIP suggesting that cognitive impairment in psychotic disorders is not simply a consequence of diagnostic category, but likely reflects an accumulation of transdiagnostic exposures, including childhood adversity, substance use, and antipsychotic prescription^2^. Building on this approach, we demonstrate that receptor-modulation profiles carry cognition-relevant information not captured by diagnostic groups or conventional medication summaries. This interpretation is supported by receptor-affinity taxonomy work showing that antipsychotic receptor profiles map onto efficacy and side-effect differences more effectively than traditional classification schemes^17^. By contrast, network meta-analytic evidence has not established a clear cognitive hierarchy among individual antipsychotic medications^4^, suggesting that drug-level comparisons may be too coarse to capture receptor-level effects on cognition.

The localization of pharmacological attribution was also notable. Although antipsychotic efficacy is closely linked to D2 receptor blockade^18^, excessive D2 occupancy has been associated with poorer neurocognitive performance^19^. Experimental studies in healthy volunteers have suggested that both D2/D3 antagonism and partial agonism can impair working memory^20^, however, previous findings across cognitive domains have been inconsistent. One possible reason is that many pharmacological challenge studies use antipsychotics with broad receptor-binding profiles^21,22^, making it difficult to attribute cognitive effects to dopaminergic blockade alone. In our analysis, D2 and D3 modulation were captured in Cluster 1, which showed a negative relationship with predicted cognition, however, this did not survive FDR correction following permutation testing.

The strongest cluster-specific signal localized to Cluster 2, spanning ADRA2A, DRD4, DRD5, HRH2, HTR1D, HTR1E, HTR2C, HTR3A and HTR6. Modulation of this cluster was associated with several antipsychotics, including aripiprazole, clozapine and olanzapine. α2A-adrenergic signaling is central to prefrontal cognitive processes^23^, while serotonergic modulation at HTR2C and HTR3 has been implicated in inhibitory control^24^, sensory processing^25^ and procognitive pharmacology. The inclusion of DRD4 is noteworthy. D4 receptors are enriched in prefrontal circuits, modulating GABAergic inhibition onto pyramidal neurons^26^. Excitatory-inhibitory (E/I) balance is thought to be crucial for numerous cognitive operations, with its disruption widely implicated in neuropsychiatric disorders^27,28^. D4 receptors, therefore, appear to hold an important role in maintaining this balance. In rodent models of schizophrenia, D4 agonism has been shown to normalize E/I abnormalities^29^, raising the question as to whether D4 agonists could improve E/I balance and cognitive function in humans.

Increased Cluster 7 modulation, corresponding to SLC6A3 activity, was positively associated with predicted cognition across models. SLC6A3 encodes the dopamine transporter, a key target of stimulant and catecholaminergic medications^30^. SLC6A3 modulation was driven predominantly by bupropion and sertraline, as well as contributions from duloxetine and methylphenidate. Several of these medications have previously been investigated with regard to cognitive modulation, though their effects have varied substantially^31,32^. Relative to broader receptor clusters, SLC6A3 was more sparsely modulated, and as such the signal should be interpreted with some caution, in part as there may be residual confounding not fully accounted for in our analyses controlling for symptom severity and diagnosis. Cluster 4 (HTR1B and HTR5A) also showed significant cluster-level attribution to composite cognition in the primary and PANSS-adjusted models, with a domain-specific negative association with verbal fluency, although the composite association did not survive FDR correction in the restricted model.

Modulation of the muscarinic Cluster 5 (CHRM1-5) was significantly associated with poorer verbal memory and motor speed. In our cohort, this cluster was driven predominantly by prescriptions of quetiapine, benztropine, and clozapine. There is substantial evidence linking greater anticholinergic burden to poorer cognition in psychosis, particularly verbal learning^7,33^. Experimental muscarinic antagonism likewise impairs memory and attentional performance^34^, while higher anticholinergic burden in schizophrenia has been associated with poorer declarative memory and complex attention, but not executive function or verbal fluency^35^. Domain-specific associations were also observed in our study, with muscarinic modulation significantly associated with poorer verbal memory and motor speed, however, there was no significant relationship with verbal fluency and executive function. Taken together, these findings suggest that the impact of muscarinic modulation may differ substantially by cognitive domain rather than reflecting a uniform deficit.

Cognitive impairment remains a major unmet need for individuals with psychotic disorders and is inadequately addressed by existing treatments^1^. Clinicians have historically faced a difficult trade-off: dopamine-blocking antipsychotics are effective for positive psychotic symptoms but may have adverse effects on cognition. Although cognitive impairment may be less overt than hallucinations or delusions, it is a stronger determinant of long-term functional outcome^1^. Treatment optimization is therefore particularly challenging for individuals in whom cognitive impairment is prominent. Recent pharmacological advances may begin to change this landscape. Xanomeline-trospium, a muscarinic agonist combination, has become the first non-dopaminergic antipsychotic licensed for schizophrenia, with secondary analyses suggesting potential cognitive benefit in those with baseline impairment^36^. Our findings provide proof of principle that medication-derived receptor modulation profiles capture cognition-relevant heterogeneity. Longitudinal studies, ideally in first-episode cohorts, should examine cognition from before antipsychotic initiation and prospectively during treatment with a single agent, incorporating objective measures of drug exposure, adherence, and substance use. Such studies could determine whether changes in receptor-modulation profiles track cognitive change over time, while randomized or other experimental designs would be required to establish causality. Ultimately such evidence could identify individuals who may benefit from closer cognitive monitoring, medication optimization, or non-pharmacological interventions, such as cognitive remediation.

## Strengths and Limitations

Strengths of this study include the large, deeply phenotyped transdiagnostic cohort and participant-level modelling of multiple psychotropics across receptor systems, incorporating competitive binding and intrinsic activity. Cross-validated machine learning and held-out SHAP estimation reduced overfitting, while within-diagnosis FMI block permutation tested whether pharmacological profiles carried signal beyond broad diagnostic prescribing patterns. To our knowledge, this is the first study to model psychotropic exposure as a participant-level, multi-drug, multi-receptor modulation profile and test its relevance to cognition in psychosis.

Several limitations should also be considered. First, the cross-sectional design precludes causal inference and remains susceptible to confounding by indication and reverse causation. Medication choice is influenced by illness severity, previous medication exposure, response and intolerance, comorbidity, adherence, and in some cases cognition itself. It can also reflect mental health services themselves, with clinician preference, geographical and formulary availability all possibly confounding associations between medication and cognition. Current medication records were not objectively verified for adherence, and previous psychotropic exposure was incompletely captured, limiting assessment of cumulative treatment effects. Equally, FMI provides an estimated rather than directly measured index of receptor modulation. Estimates were derived from recorded dose, literature-derived receptor affinity and intrinsic activity values, and pharmacokinetic assumptions, so cannot fully capture individual differences in metabolism, blood-brain penetration, receptor availability, or endogenous ligand competition. Genetically influenced pharmacokinetic variability was likewise not incorporated, whilst affinity estimates were derived across heterogeneous experimental systems that may not directly reflect in vivo human target affinity. Furthermore, we did not model the pharmacodynamic impact of drug metabolites, which are clinically important for several compounds, as well as other potentially cognition-relevant exposures, such as nicotinic receptor modulation from tobacco use and medications lacking suitable receptor-level affinity data. Receptor clusters were also derived across the full medicated sample rather than independently within each cross-validation fold; although clustering was unsupervised and did not use cognition, the resulting feature structure was therefore shared across folds. Finally, because FMI clusters represent correlated receptor-modulation profiles, cluster-level findings should not be interpreted as isolated receptor-specific effects.

Whilst cognition in psychosis remains strongly shaped by developmental, sociodemographic, and illness-related factors, the pharmacology of psychotropic medications also appears to contribute to cognitive heterogeneity. Crucially, this signal localised to specific receptor-modulation profiles, suggesting that the cognitive effects of psychotropic medication may depend more on patterns of receptor engagement than medication class or dose alone. Modelling medications according to their receptor pharmacology may therefore provide a more informative framework for understanding treatment-related cognitive effects and, ultimately, for developing more personalised prescribing strategies.

## Data Availability

BSNIP data is available for download at https://nda.nih.gov/

## Acknowledgements

**CJW** is supported by a UKRI MRC Clinical Research Training Fellowship (UKRI3633). **MT** is supported by the NIHR as a clinical lecturer. **RU, JH, RAM** are supported by the NIHR Oxford Health Biomedical Research Centre. The views expressed are those of the author(s) and not necessarily those of the NIHR or the Department of Health and Social Care. **TP** is supported by a Wellcome Trust Early Career Award (313979/Z/24/Z), the Maudsley Charity (grant reference 3542), and the UK Research and Innovation Hub for Metabolic Psychiatry (grant reference MR/Z503563/1, platform grant code MR/Z000548/1).

## AI declaration

During the preparation of this work, OpenAI’s ChatGPT and Anthropic’s Claude were used improve the clarity and grammar of the manuscript. The author reviewed and edited all output and take full responsibility for the content of the publication.

