## Supplemental Figures for "Psychotropic Receptor Modulation and Cognition in Psychotic Disorders"

**Supplementary Figure S1. Within diagnosis permutation testing across models**

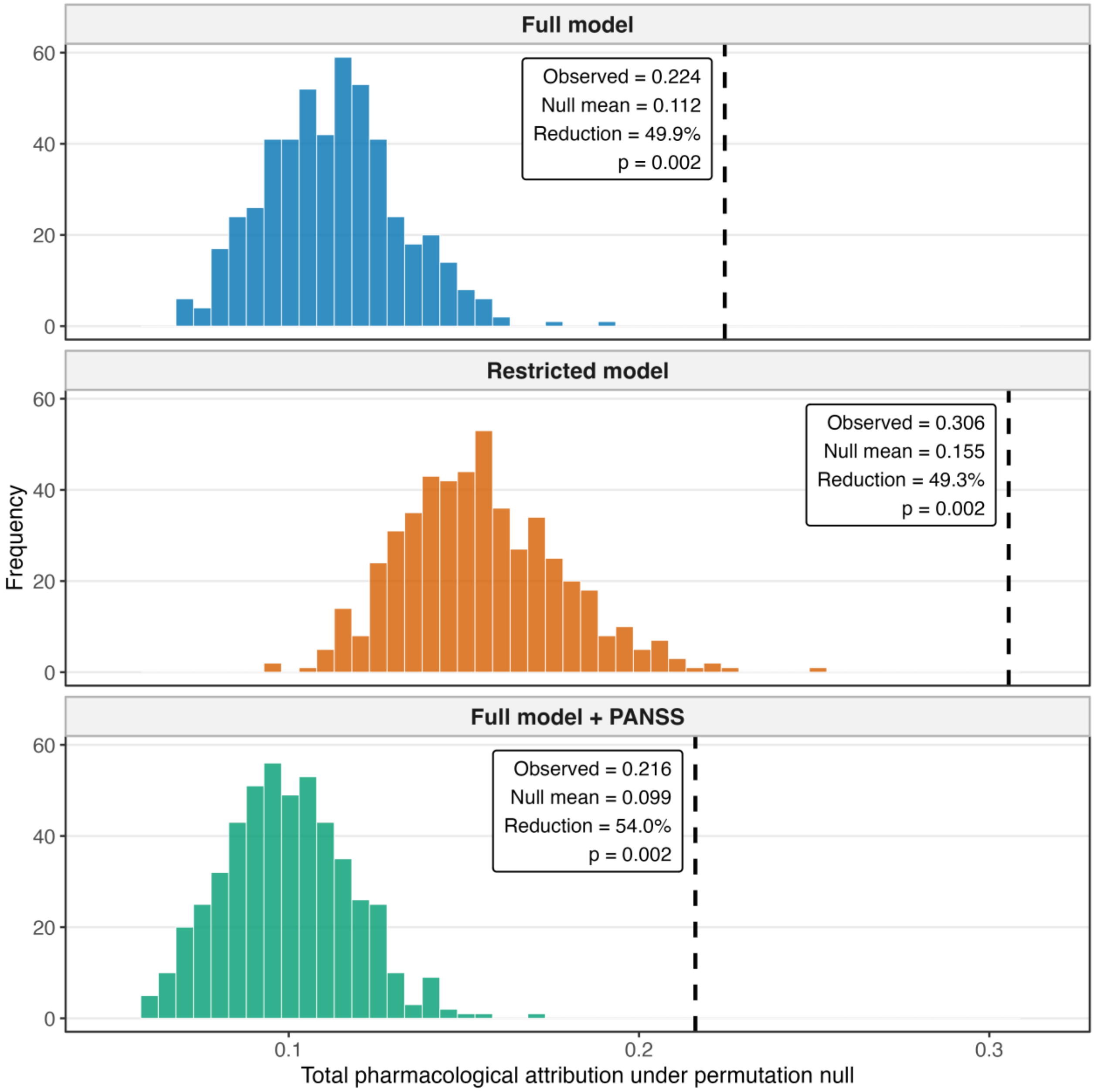

**Supplementary Figure S2. Full covariate model - diagnostics and attribution on composite cognitive scores.**

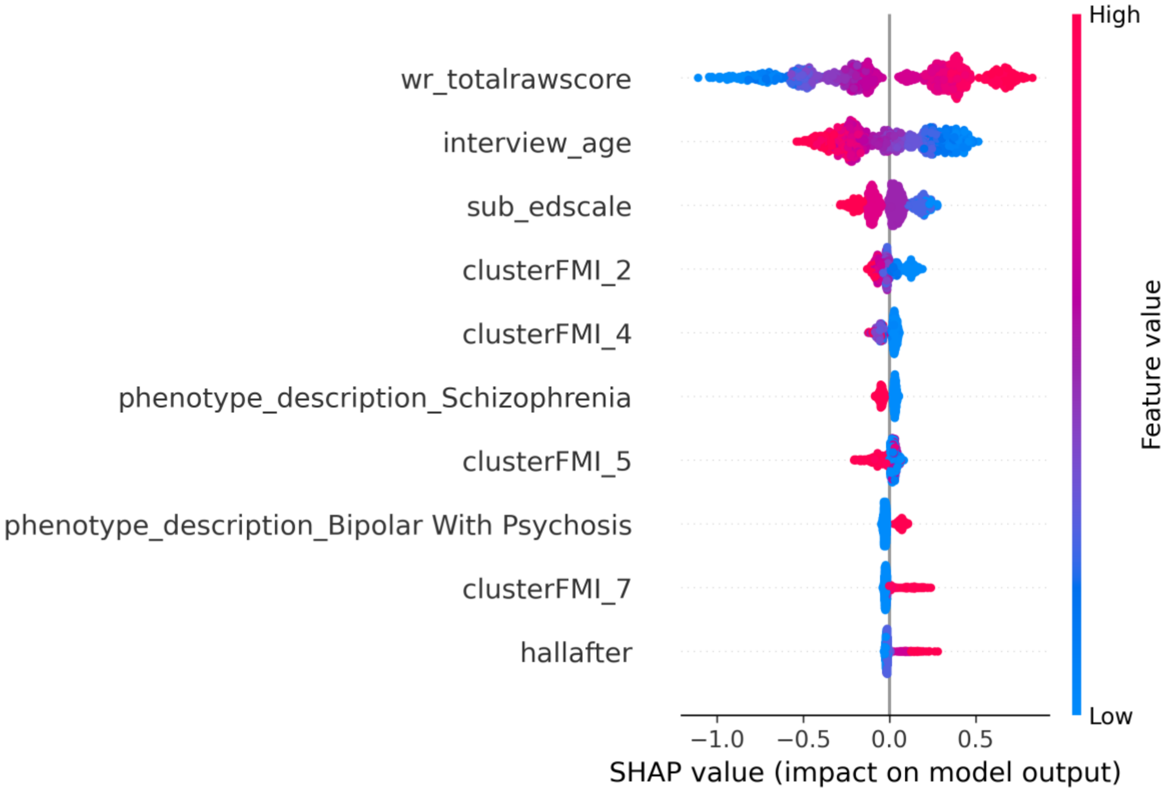


**S2A. SHAP summary plot**

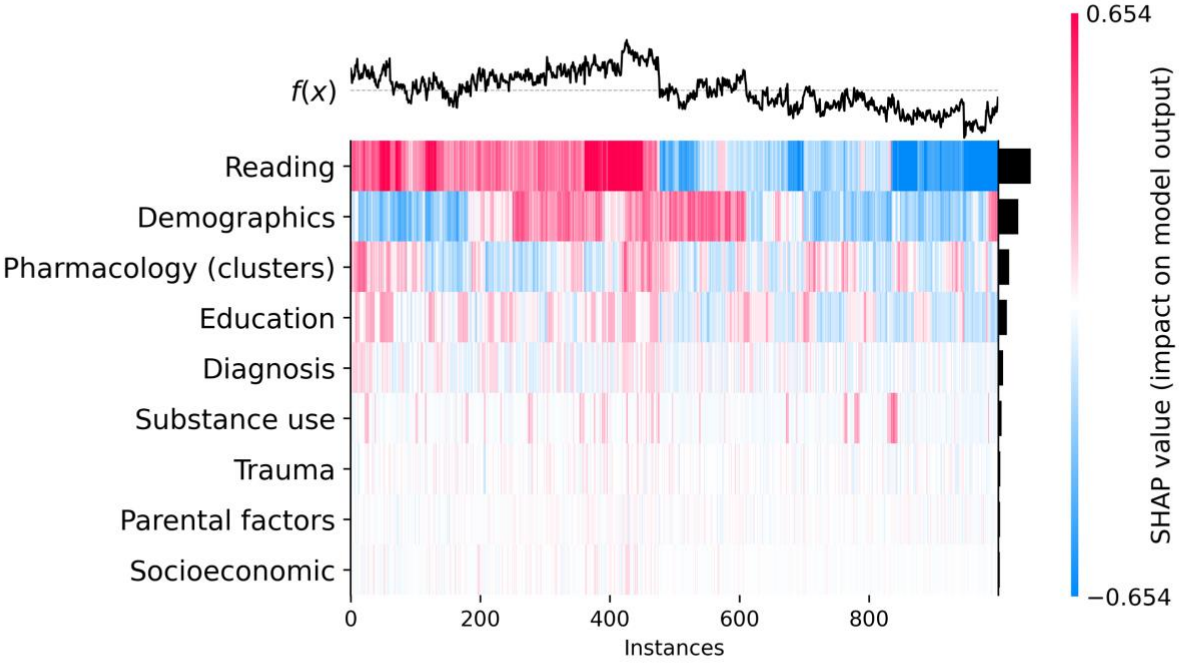


**S2B: Grouped SHAP heatmap**

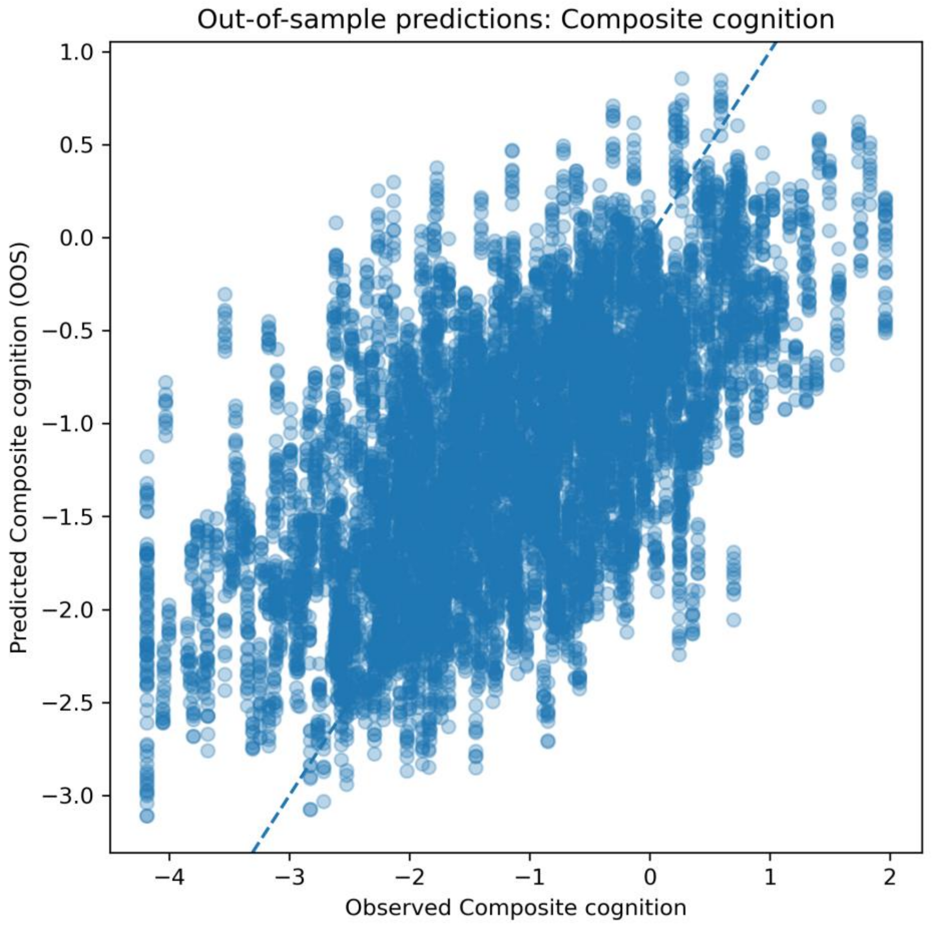


**S2C: Out-of-sample predicted versus observed cognition**

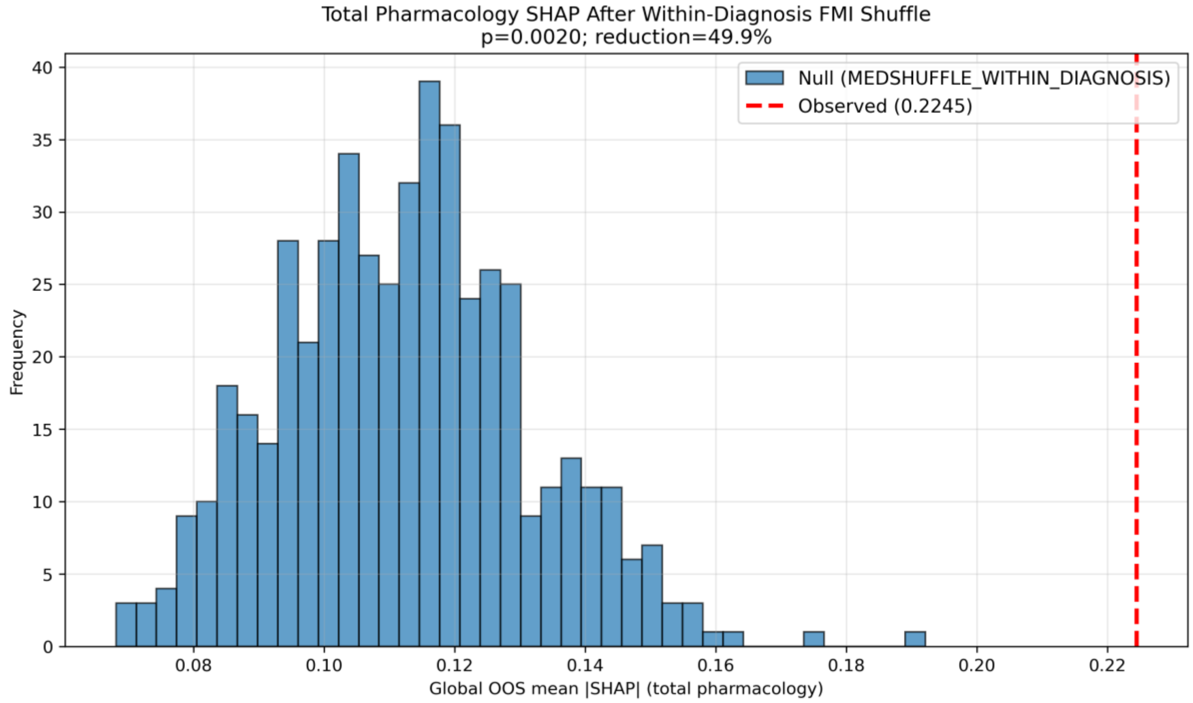


**S2D: Total pharmacology permutation**

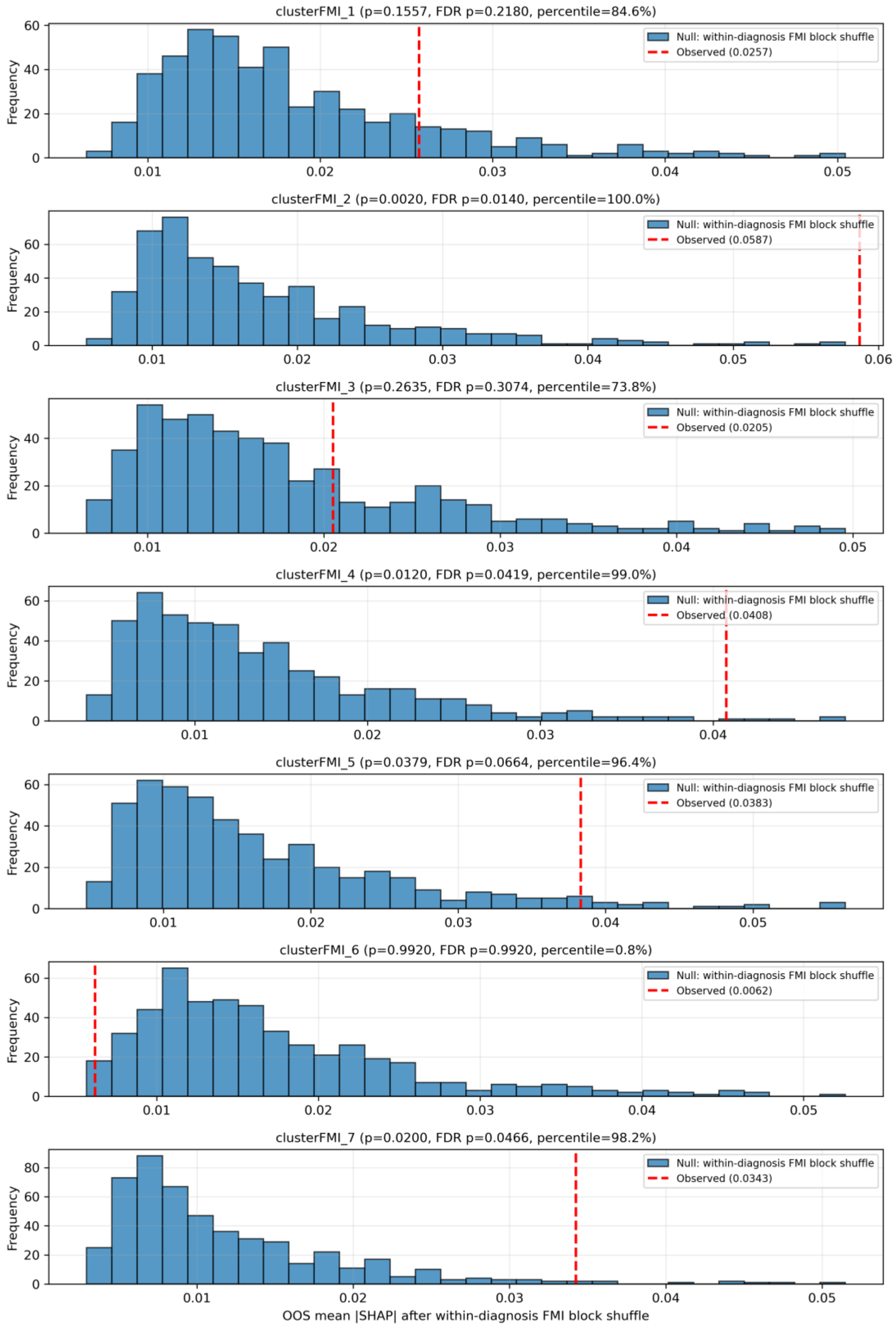


**S2E: Cluster-specific permutation null distributions**

**Supplementary Figure S3. Restricted covariate model (excluding WRAT, education and substance use) - diagnostics and attribution on composite cognitive scores.**

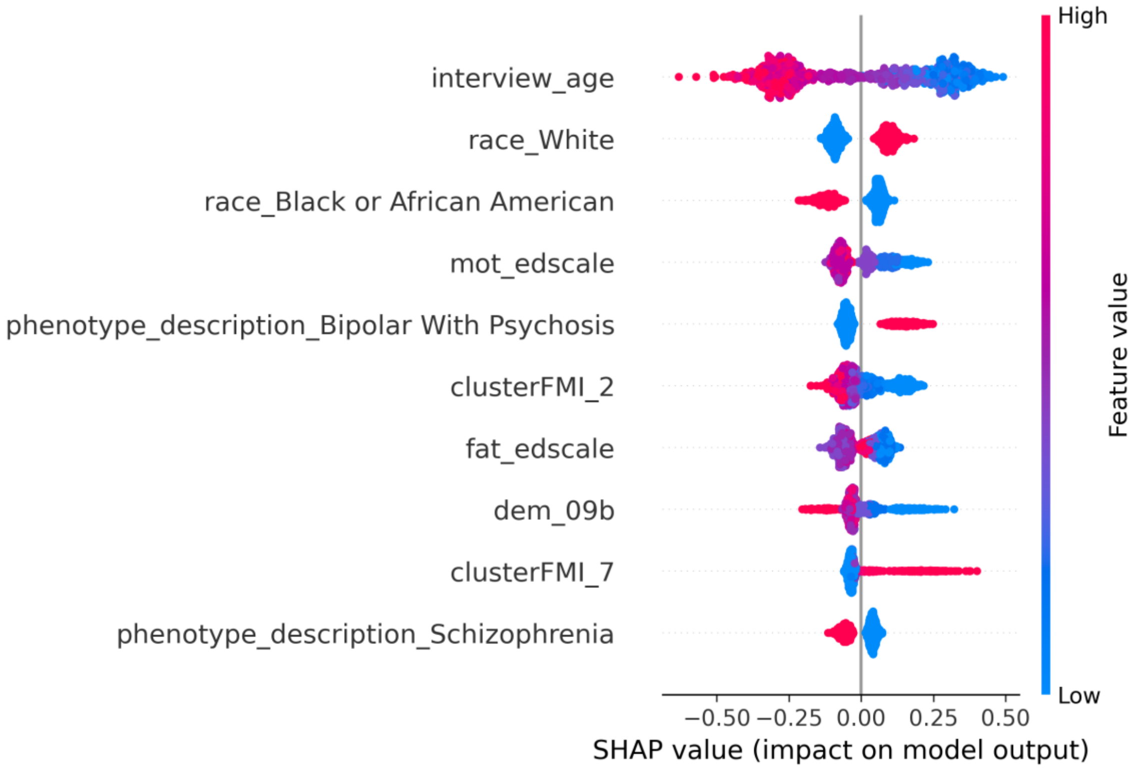


**S3A. SHAP summary plot**

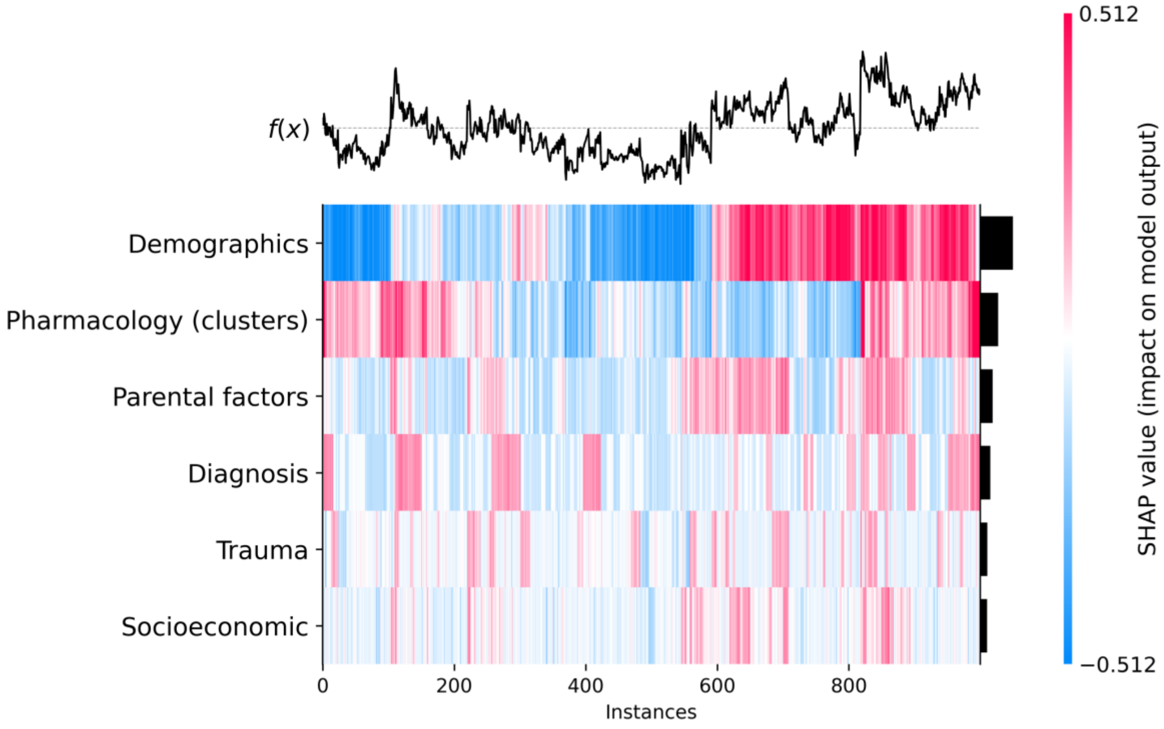


**S3B: Grouped SHAP heatmap**

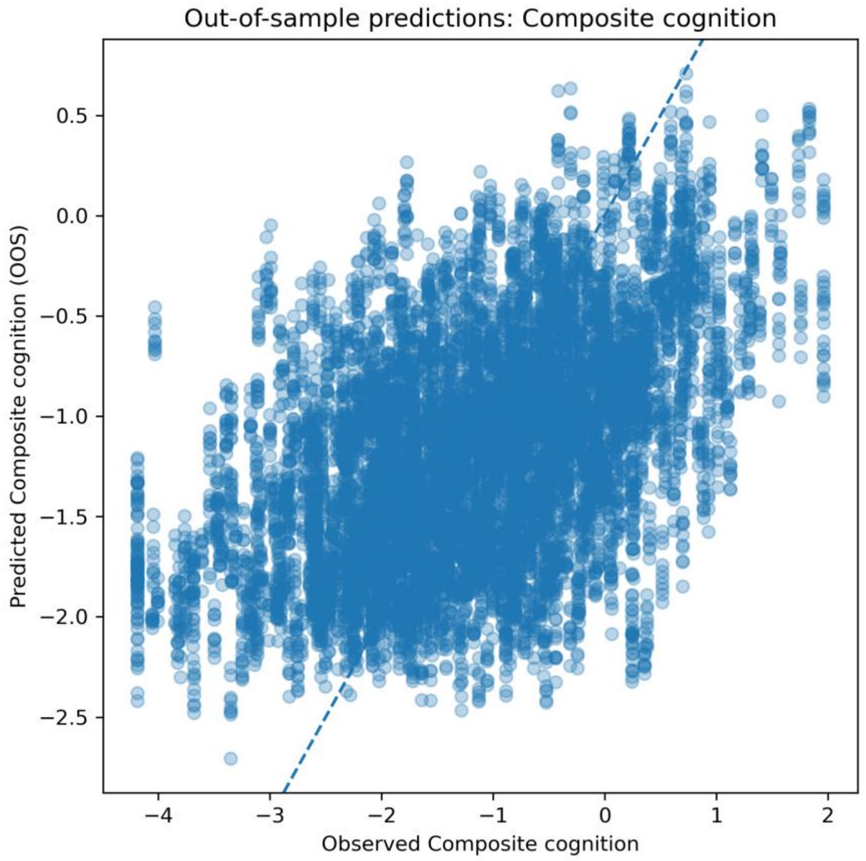


**S3C: Out-of-sample predicted versus observed cognition**

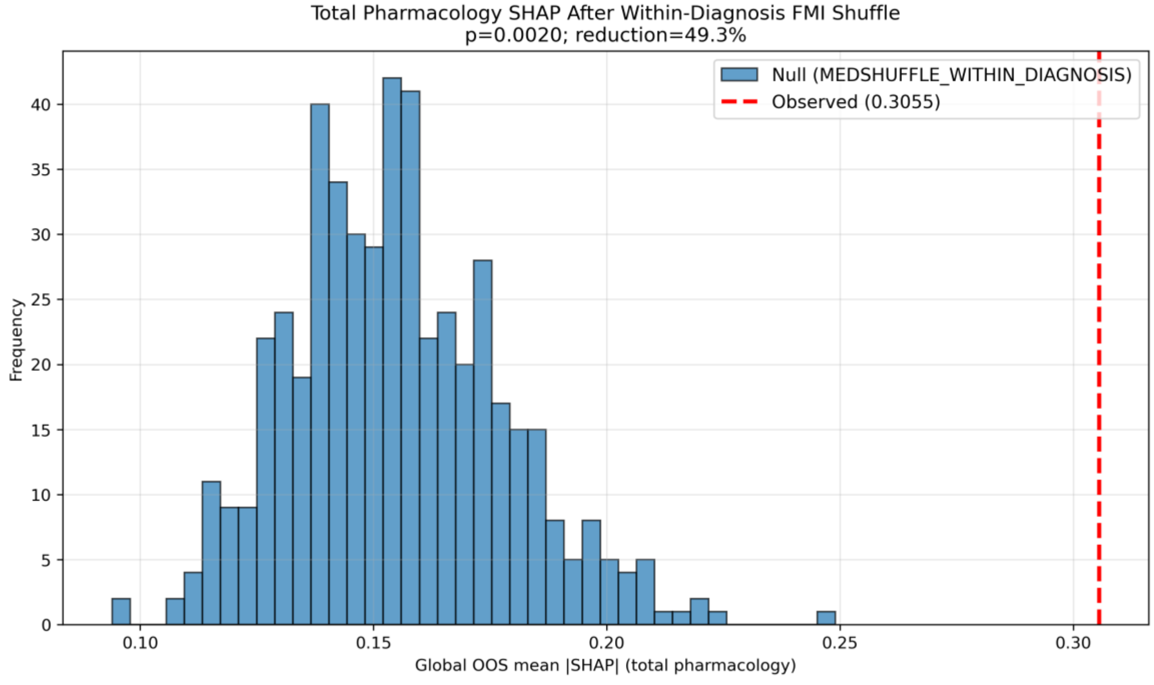


**S3D: Total pharmacology permutation**

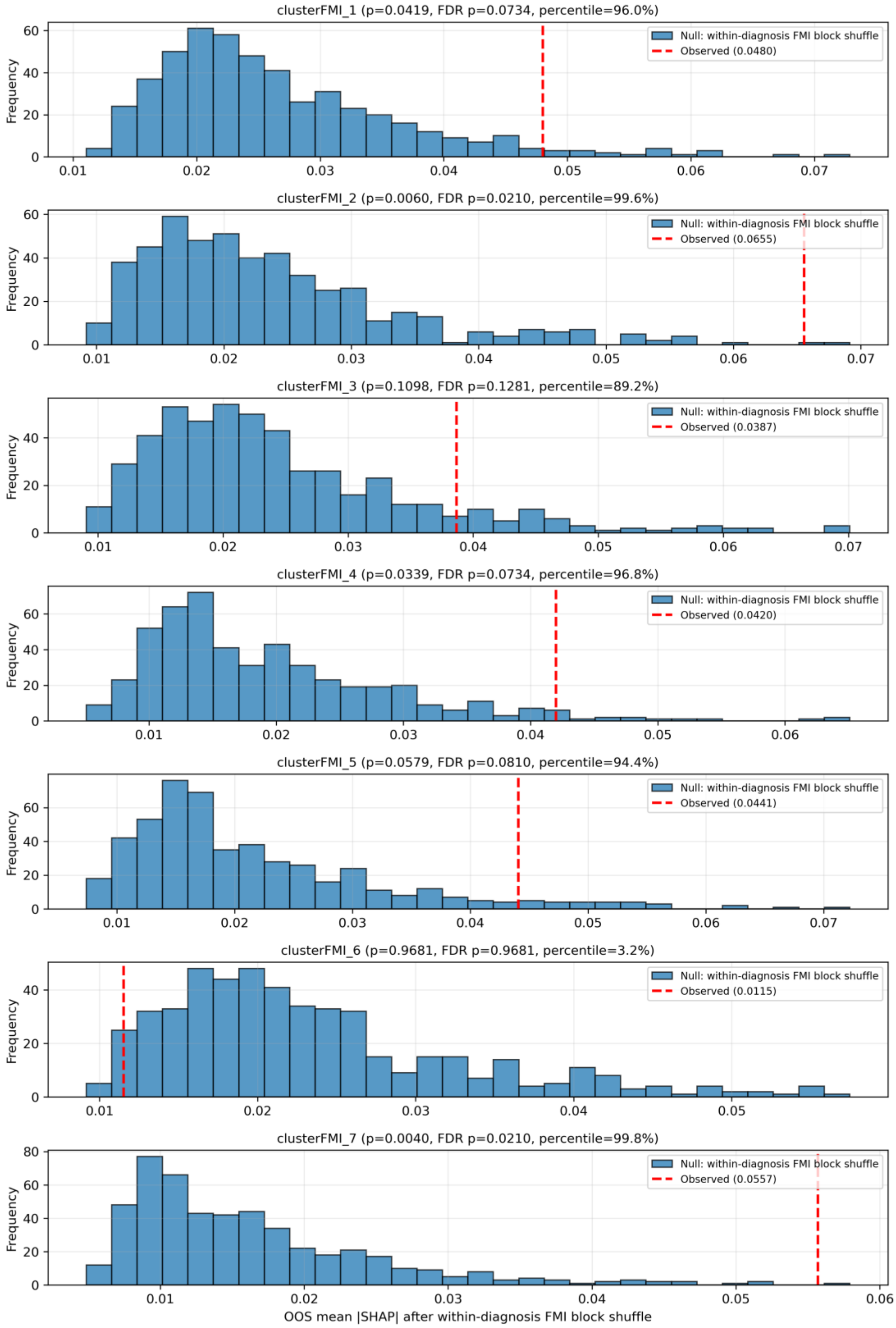


**S3E: Cluster-specific permutation null distributions**

**Supplementary Figure S4. Full covariate model plus PANSS - diagnostics and attribution on composite cognitive scores.**

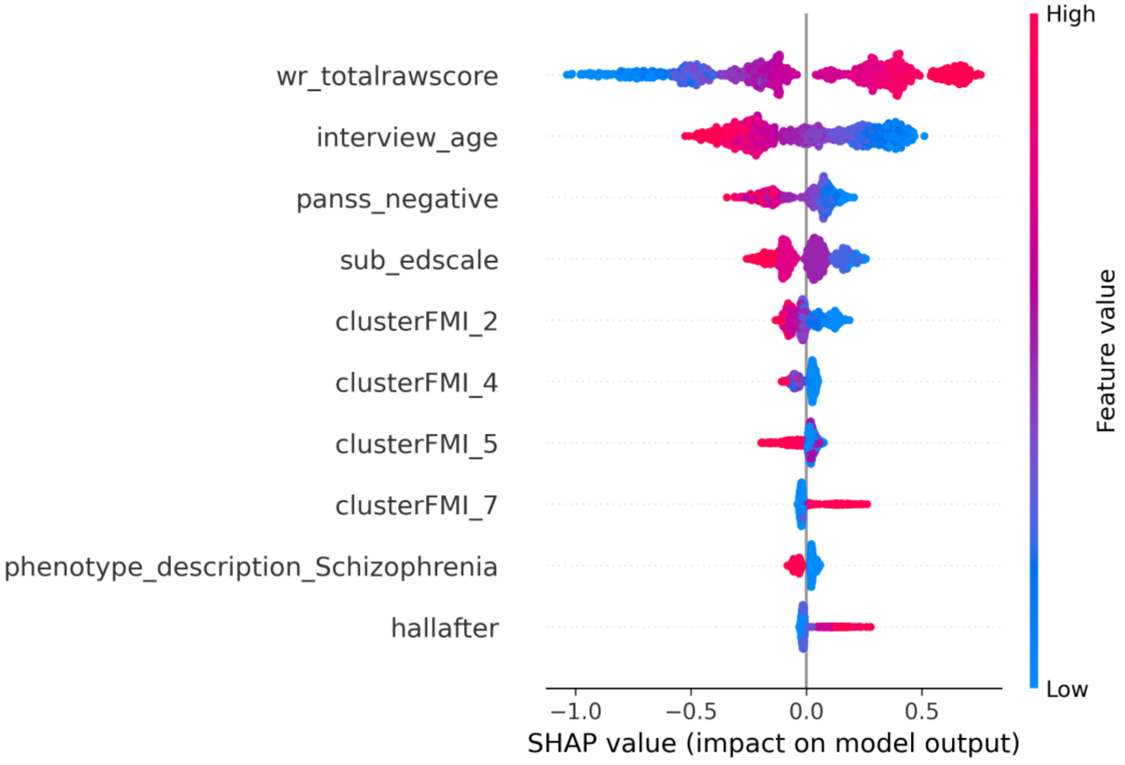


**S4A. SHAP summary plot**

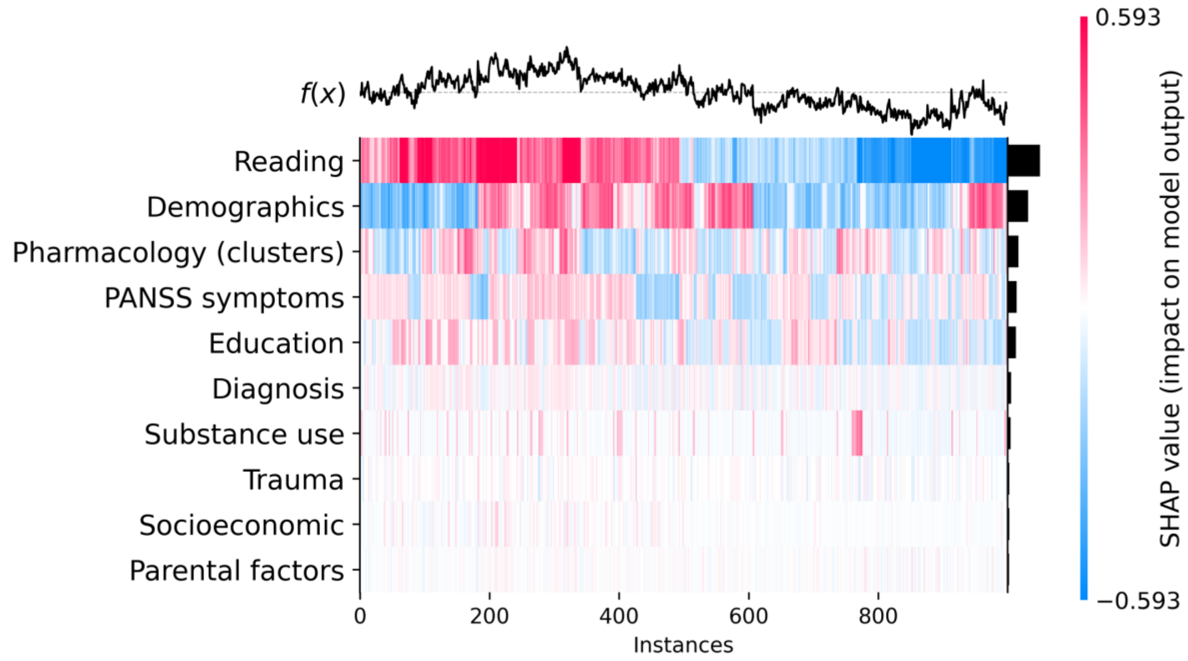


**S4B: Grouped SHAP heatmap**

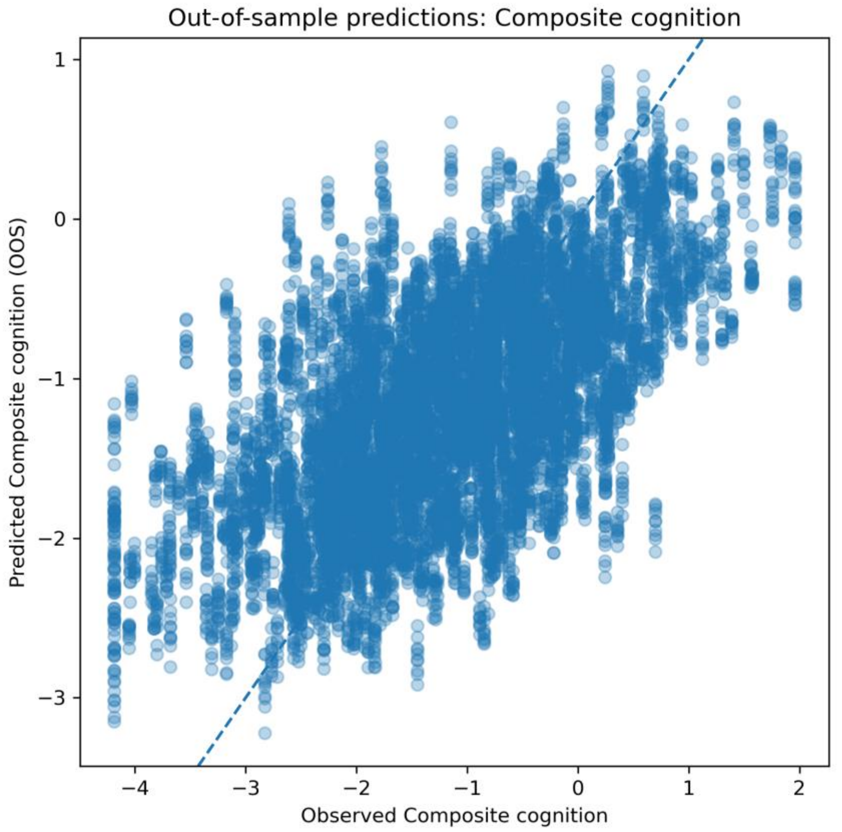


**S4C: Out-of-sample predicted versus observed cognition**

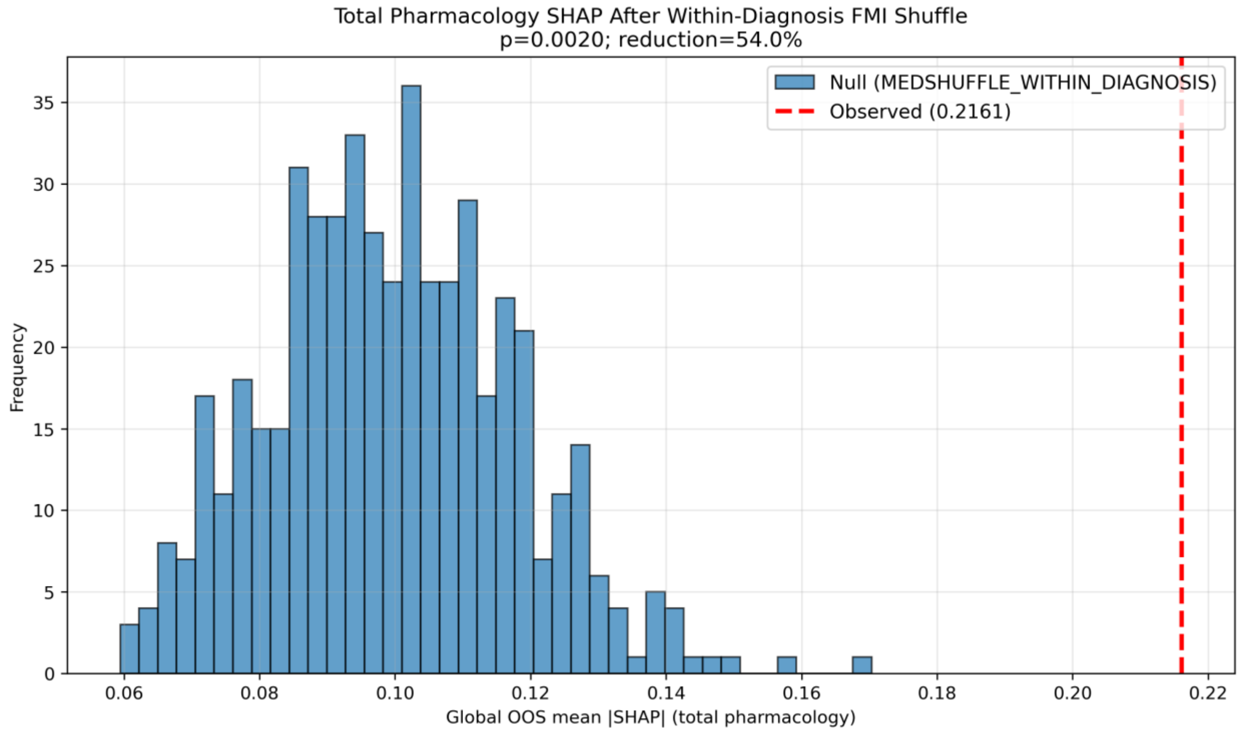


**S4D: Total pharmacology permutation**

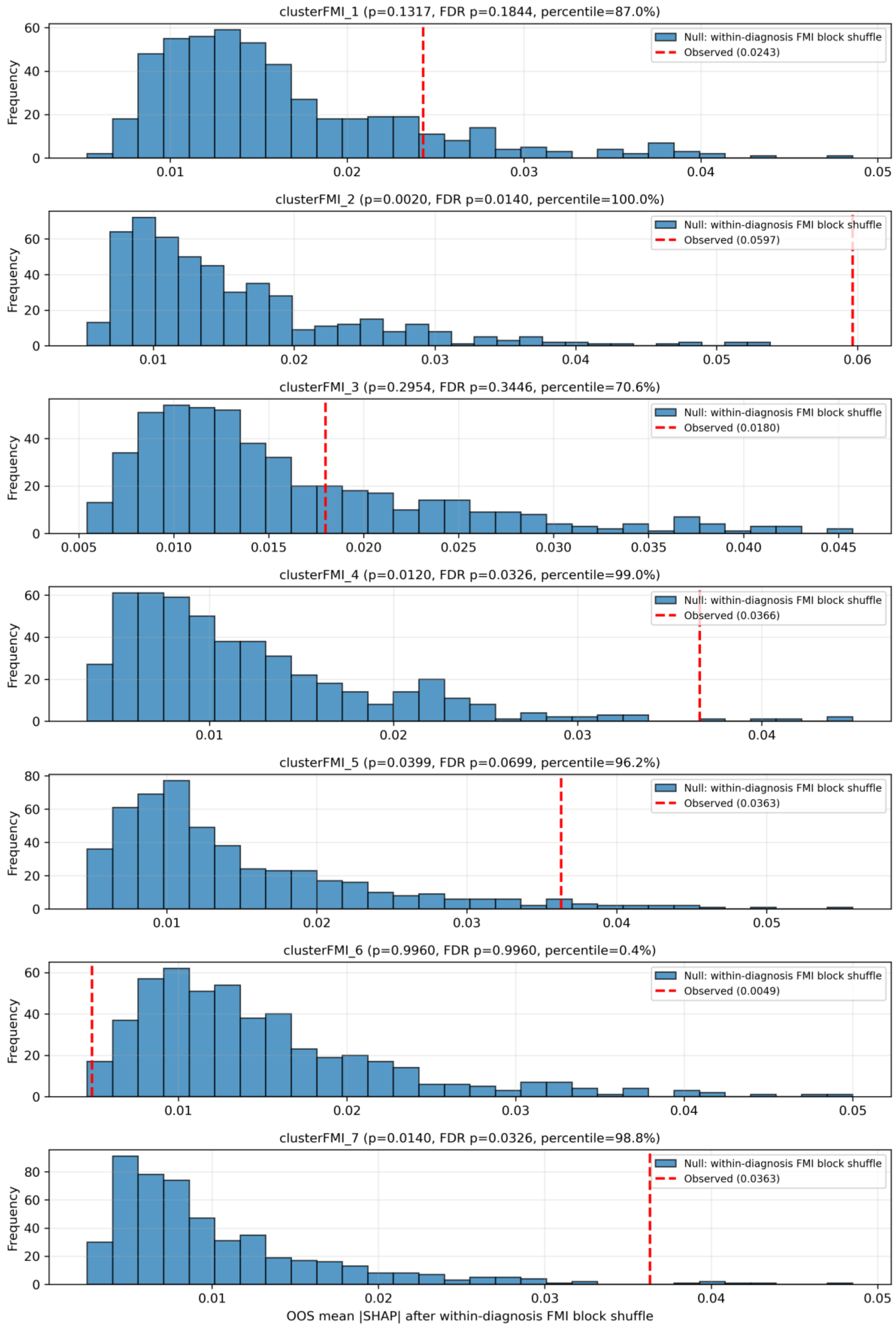


**S4E: Cluster-specific permutation null distributions**

**Supplementary Figure S5. Full covariate model - diagnostics and attribution on verbal memory scores.**

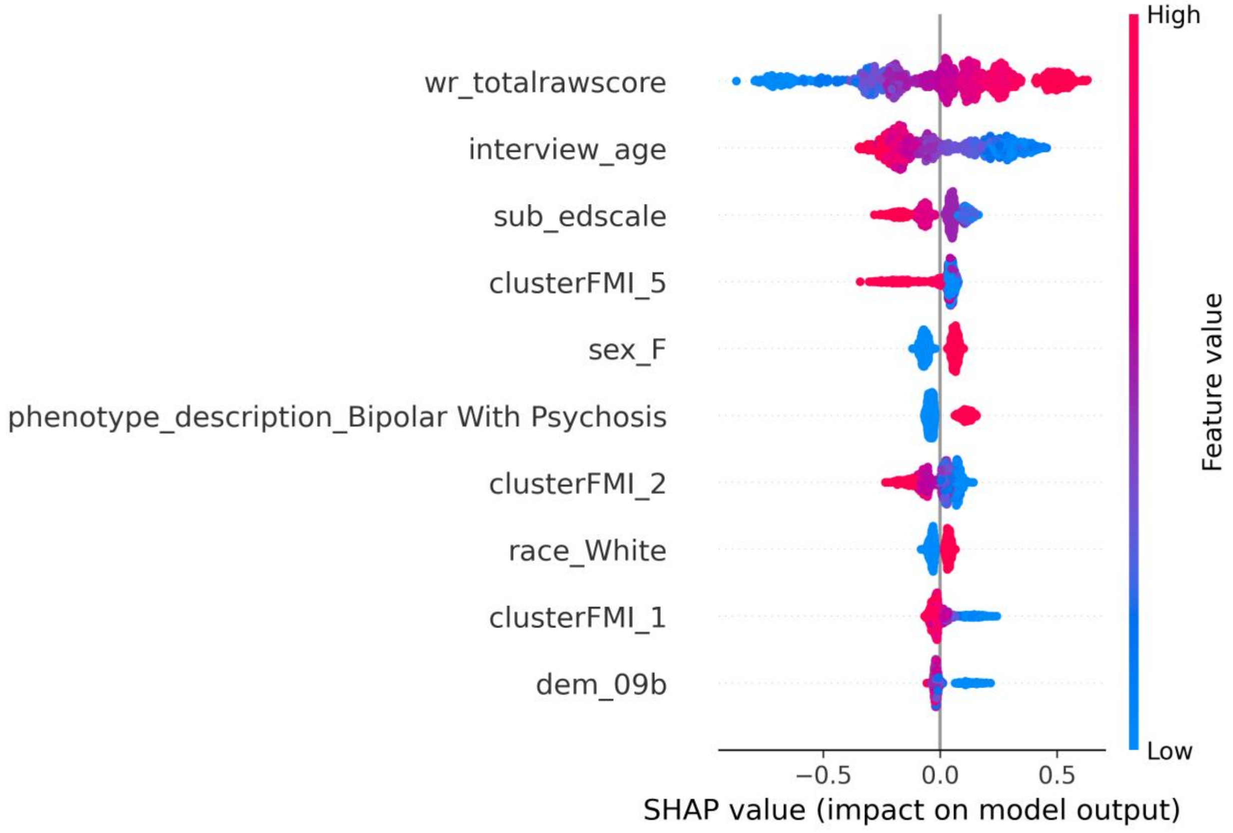


**S5A. SHAP summary plot**

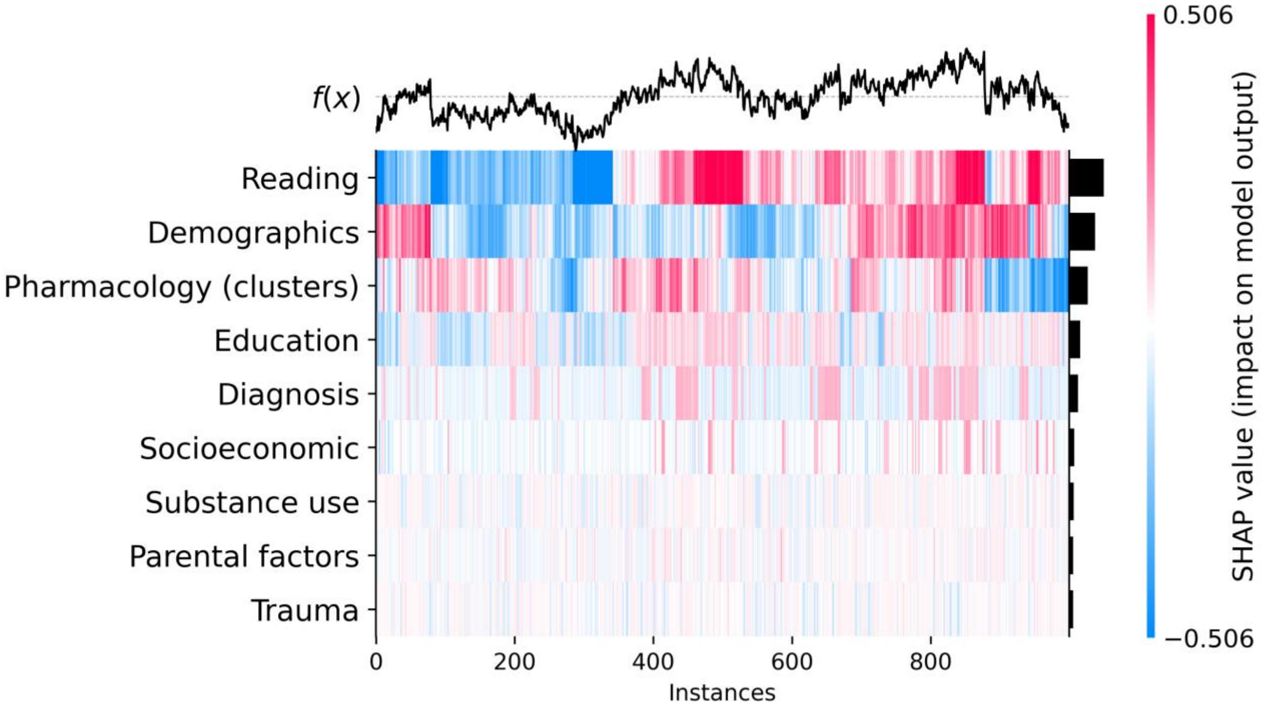


**S5B: Grouped SHAP heatmap**

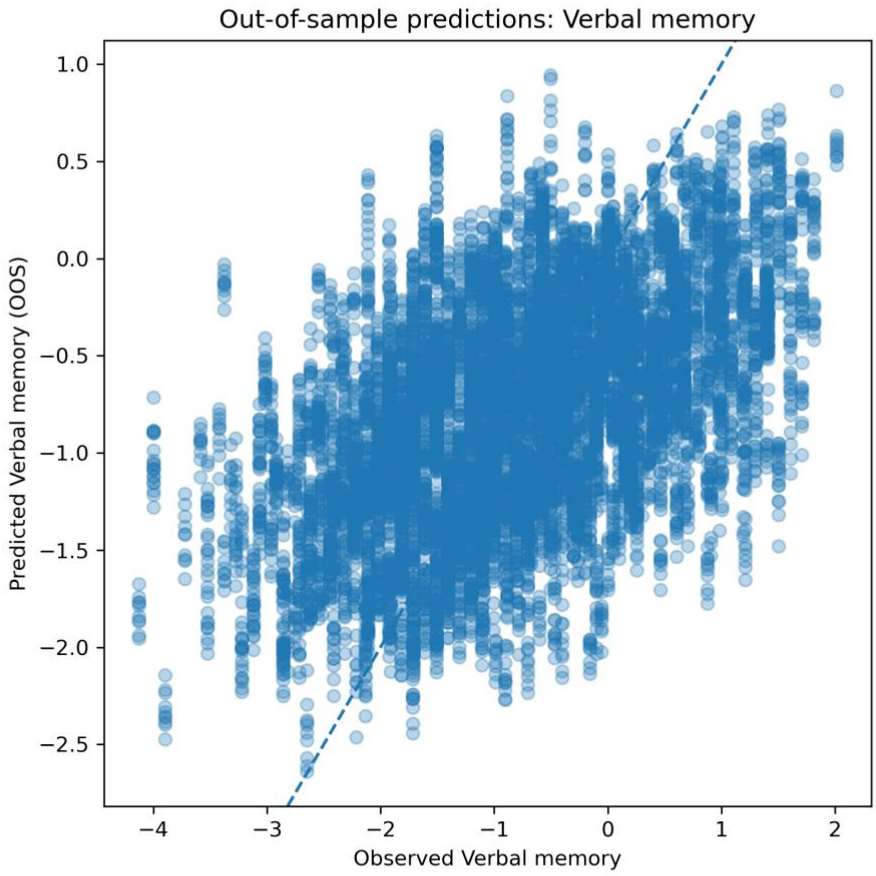


**S5C: Out-of-sample predicted versus observed verbal memory**

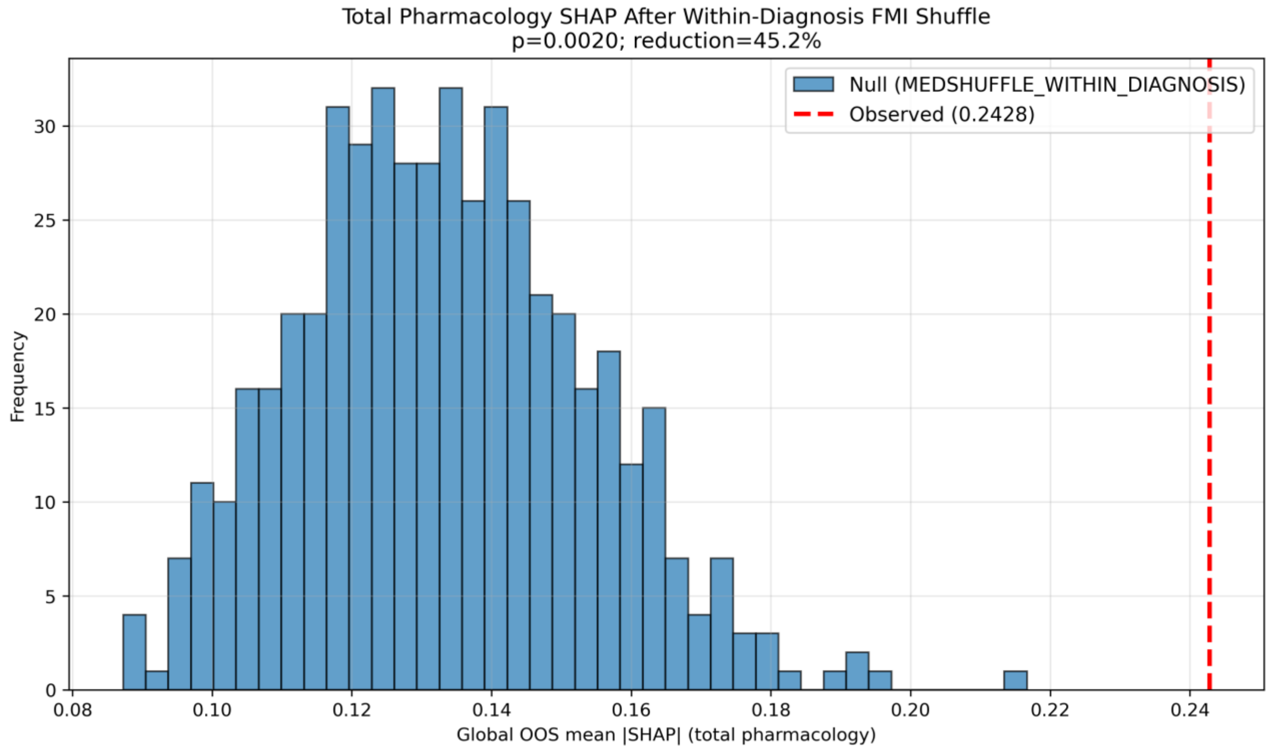


**S5D: Total pharmacology permutation**

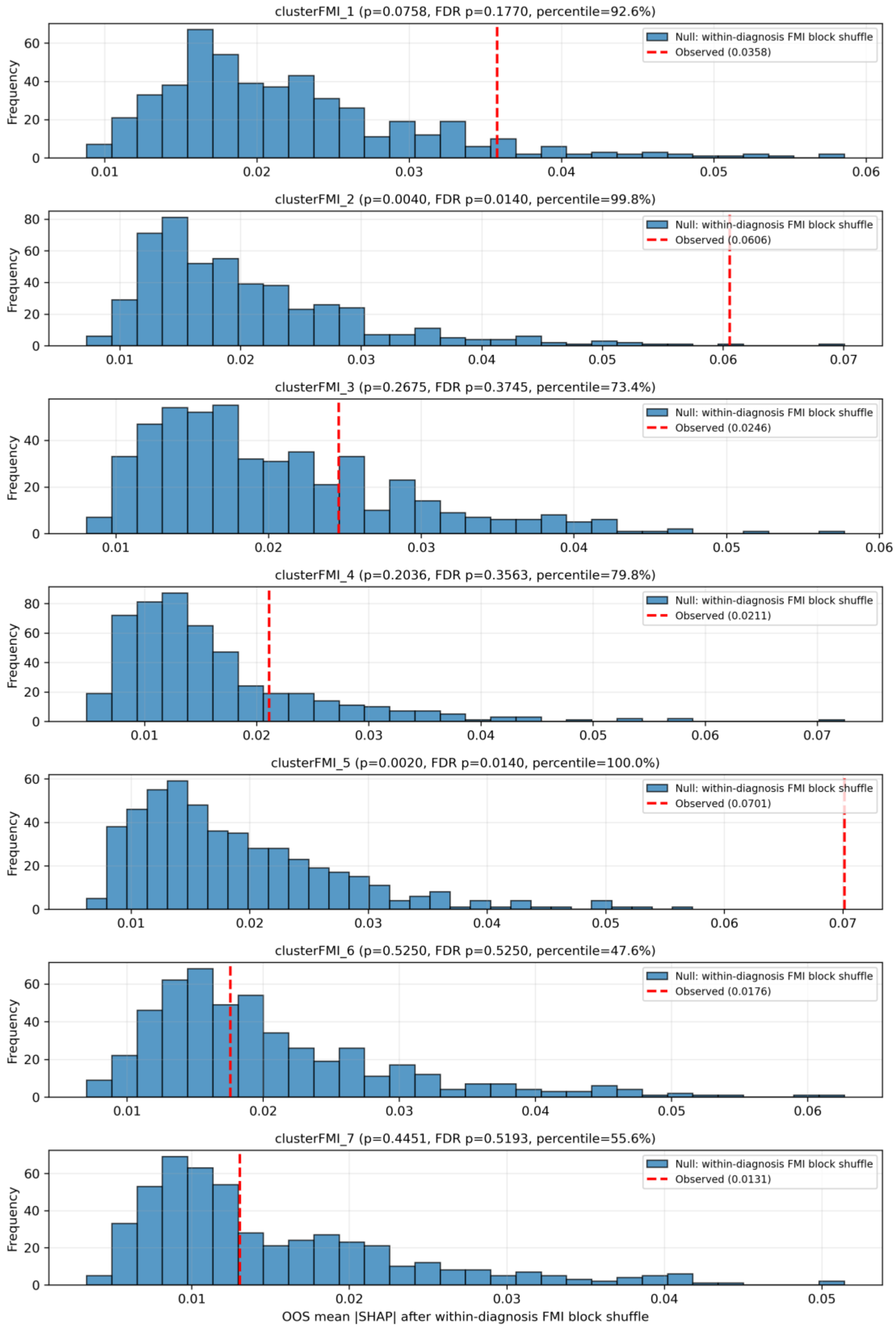


**S5E: Cluster-specific permutation null distributions**

**Supplementary Figure S6. Full covariate model - diagnostics and attribution on working memory scores.**

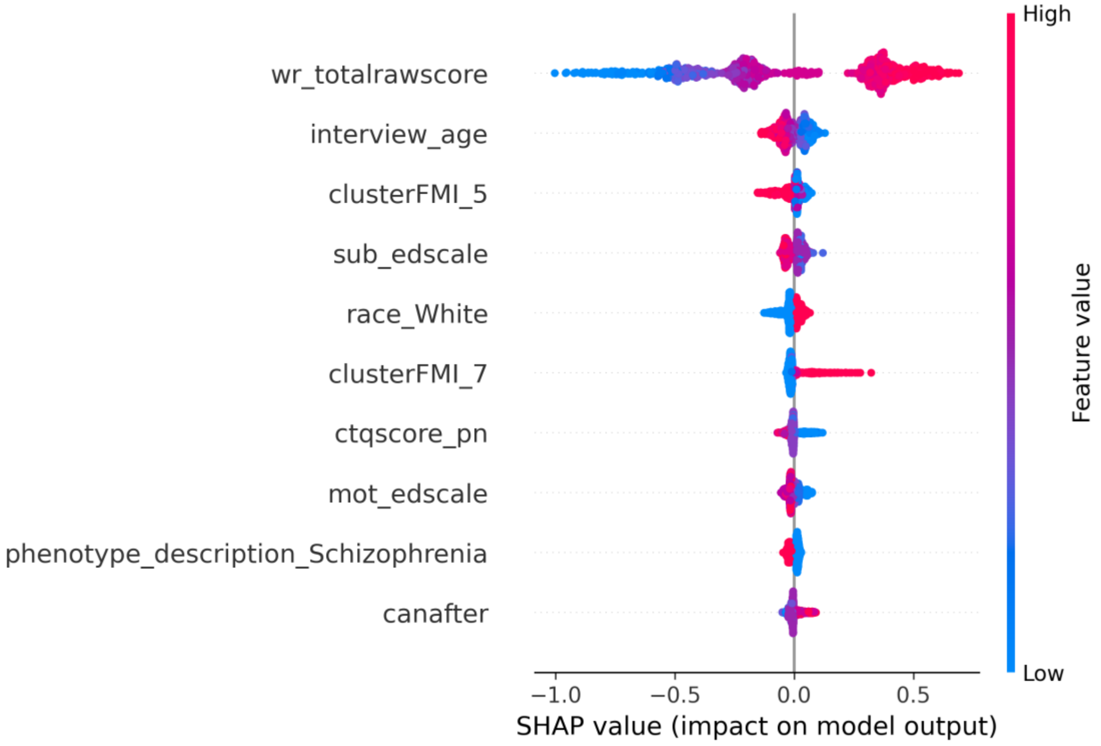


**S6A. SHAP summary plot**

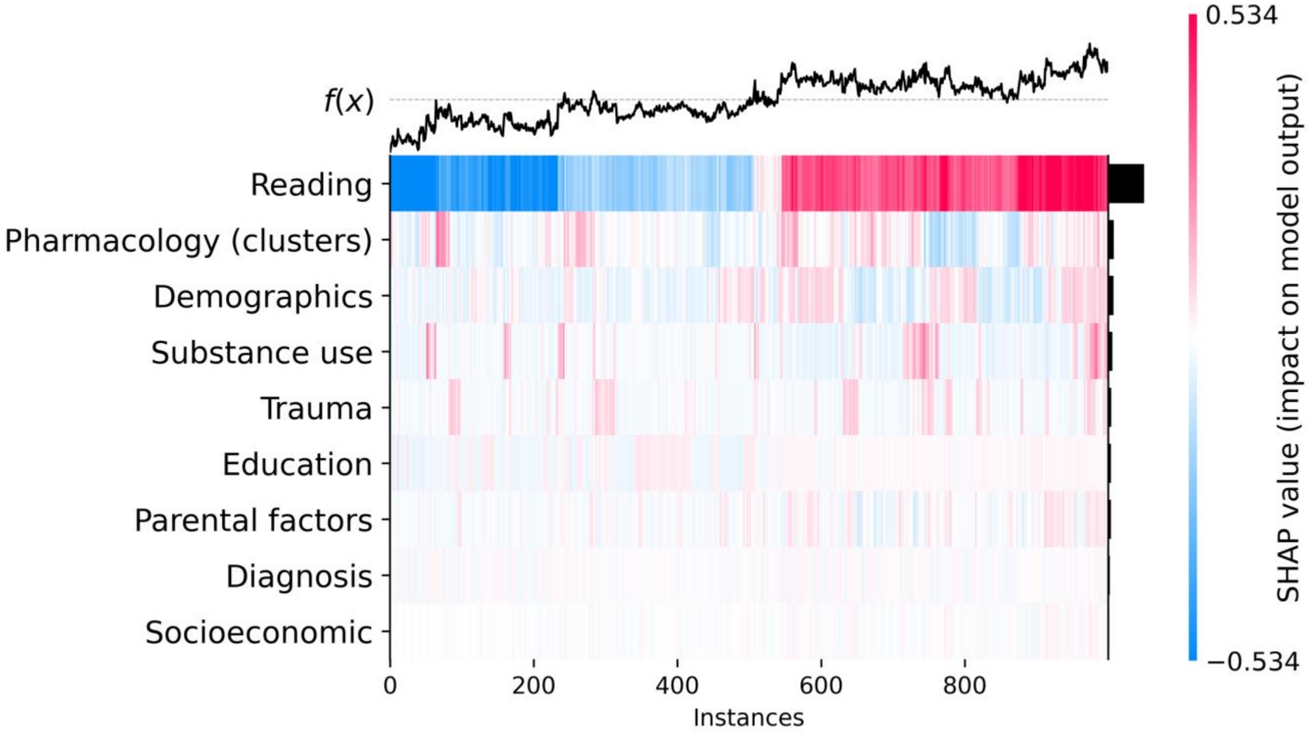


**S6B: Grouped SHAP heatmap**

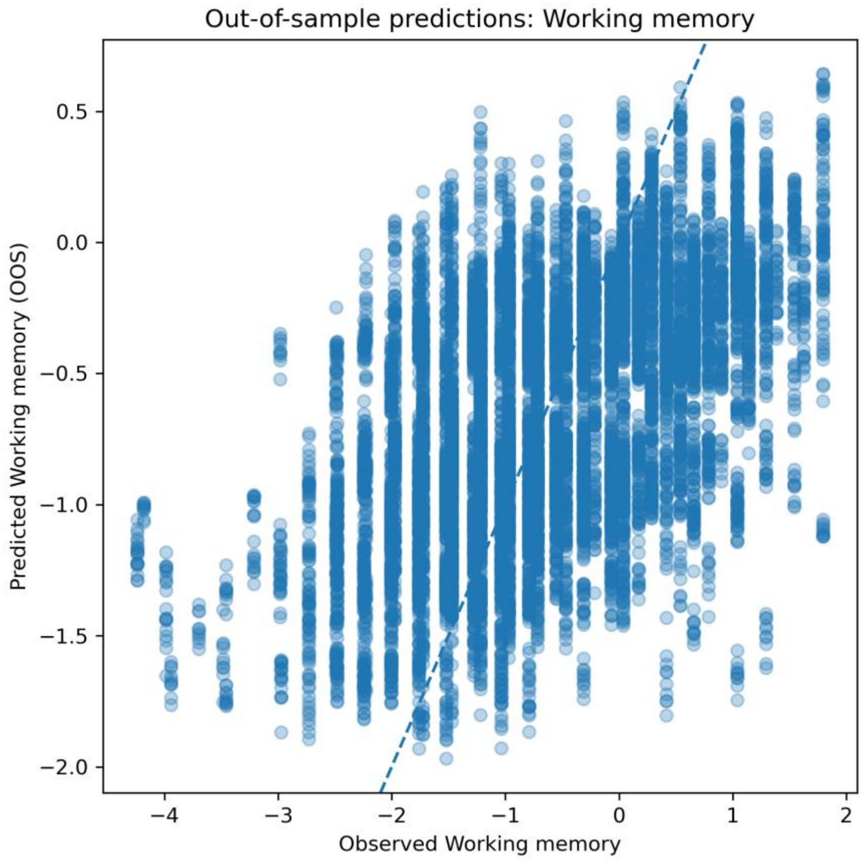


**S6C: Out-of-sample predicted versus observed working memory**

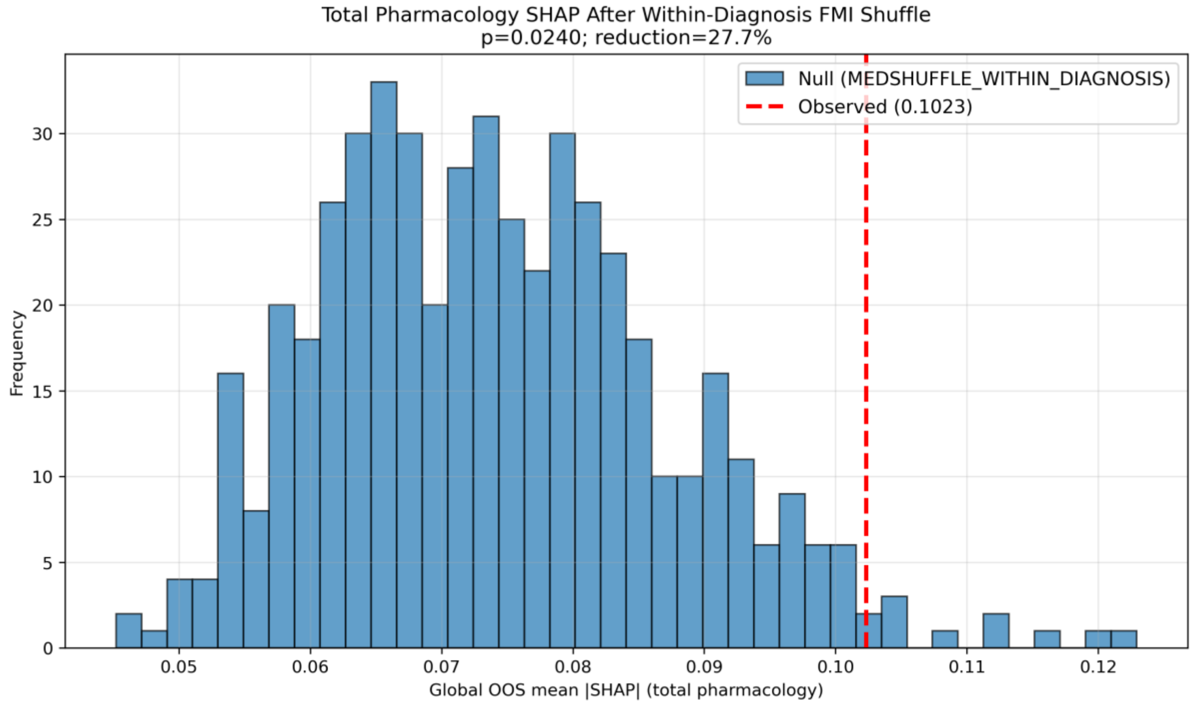


**S6D: Total pharmacology permutation**

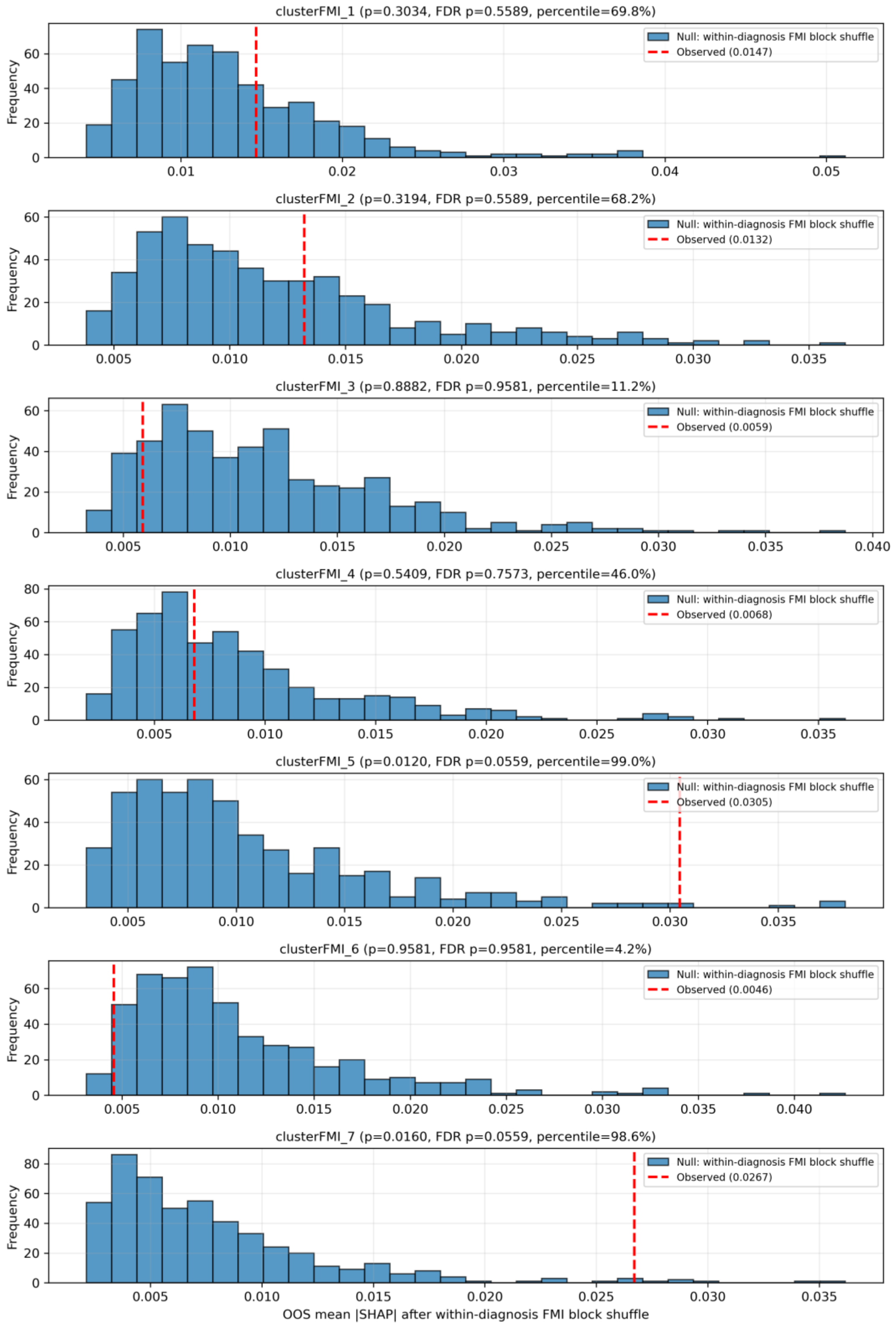


**S6E: Cluster-specific permutation null distributions**

**Supplementary Figure S7. Full covariate model - diagnostics and attribution on motor speed scores.**

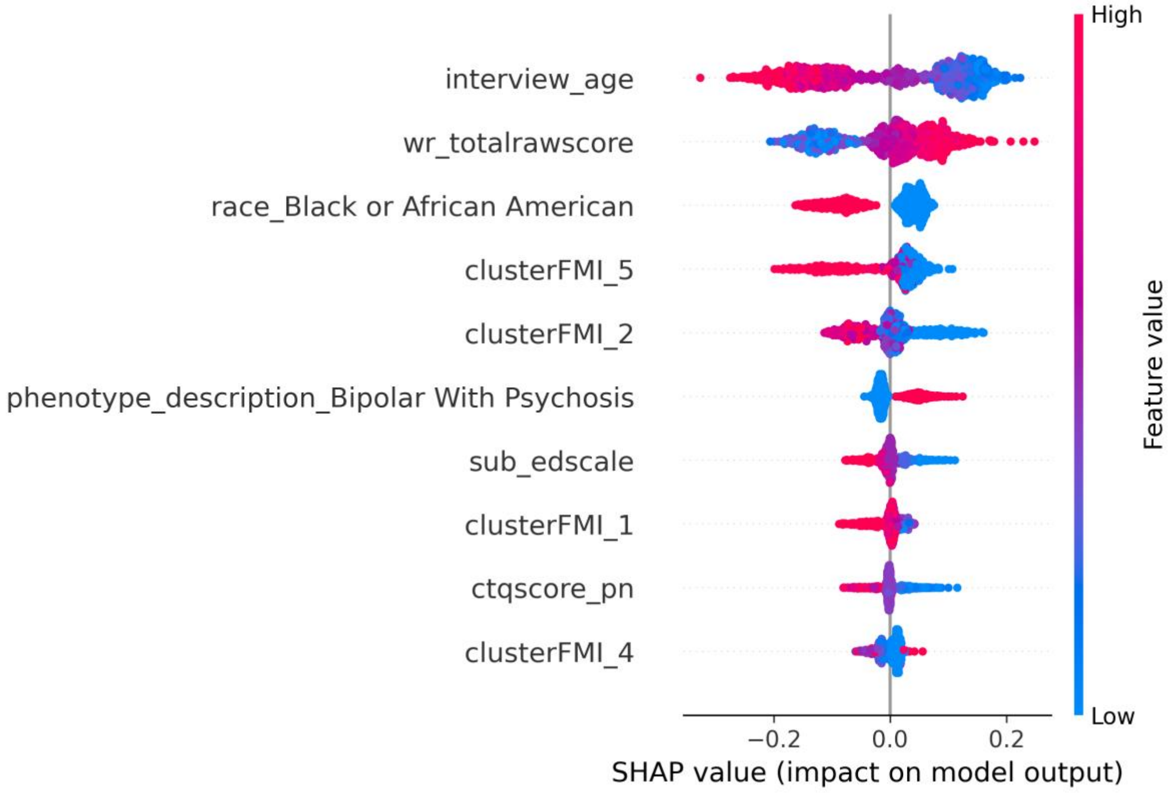


**S7A. SHAP summary plot**

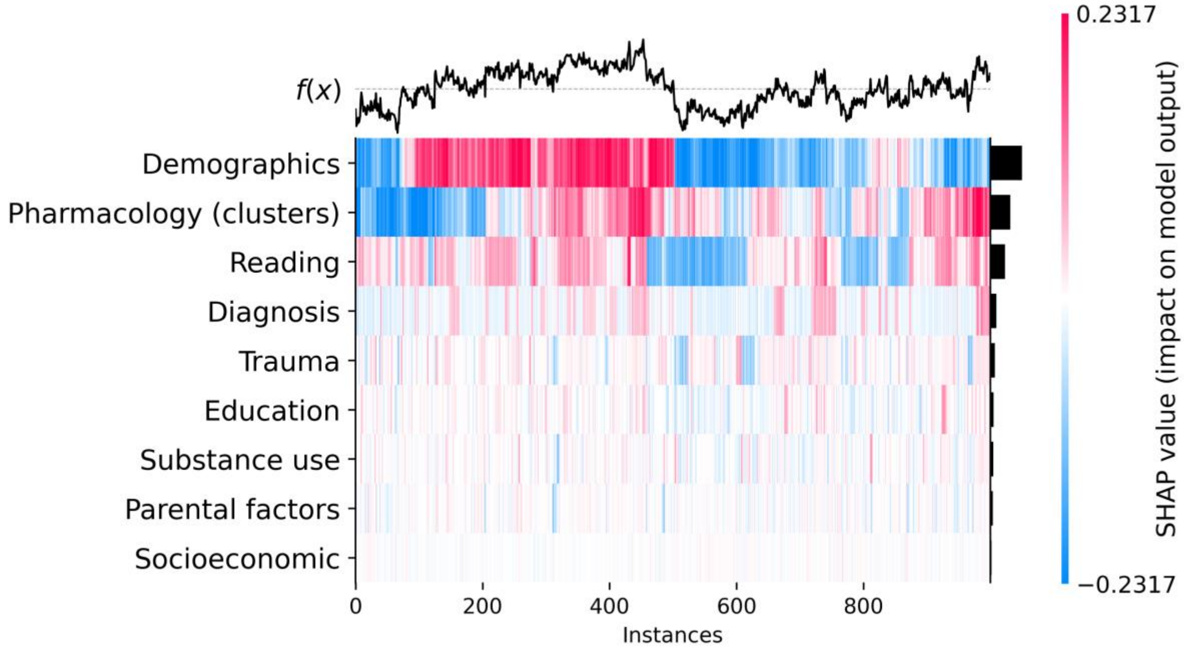


**S7B: Grouped SHAP heatmap**

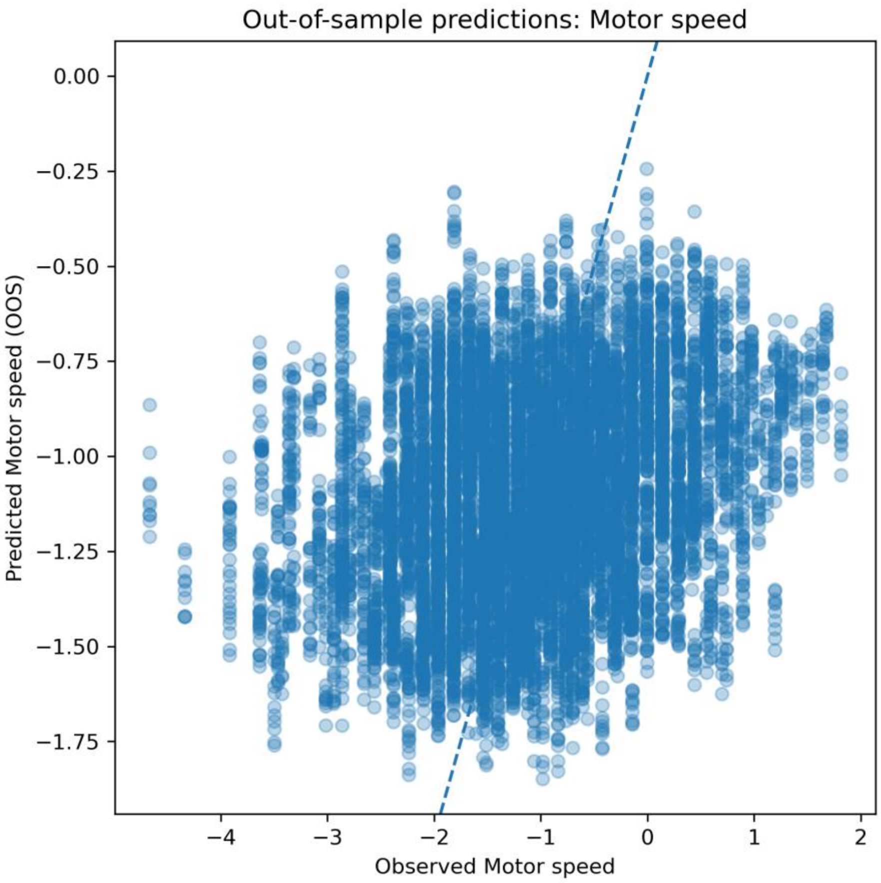


**S7C: Out-of-sample predicted versus observed motor speed**

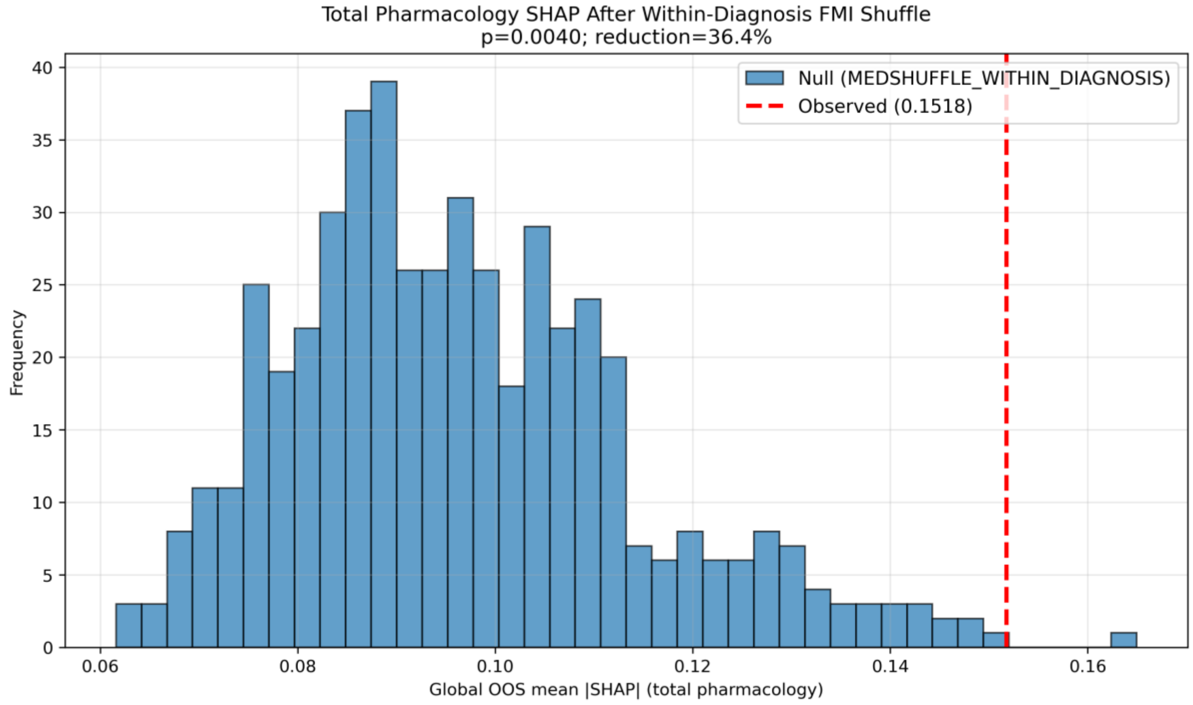


**S7D: Total pharmacology permutation**

**b**

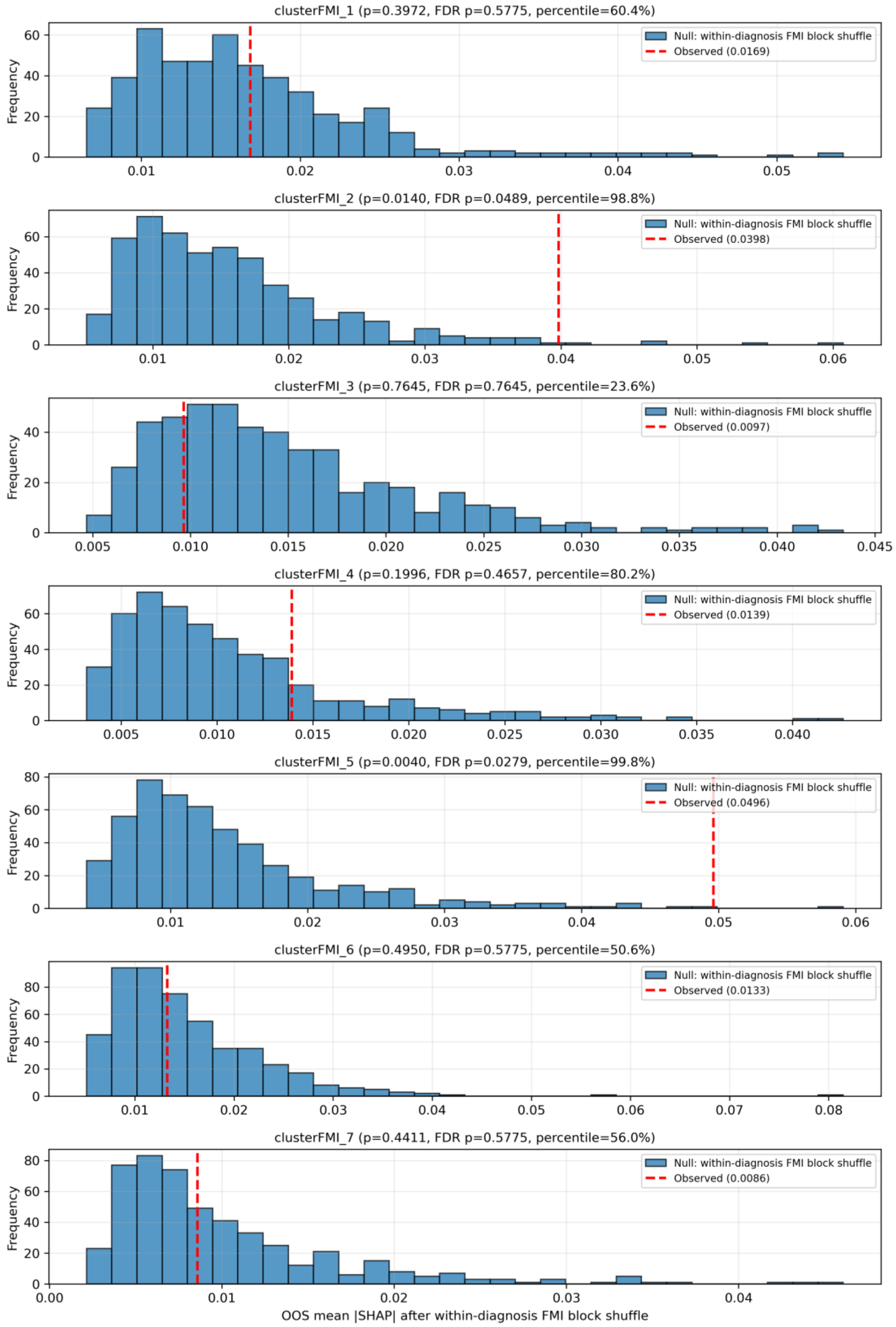


**S7E: Cluster-specific permutation null distributions**

**Supplementary Figure S8. Full covariate model - diagnostics and attribution on verbal fluency.**

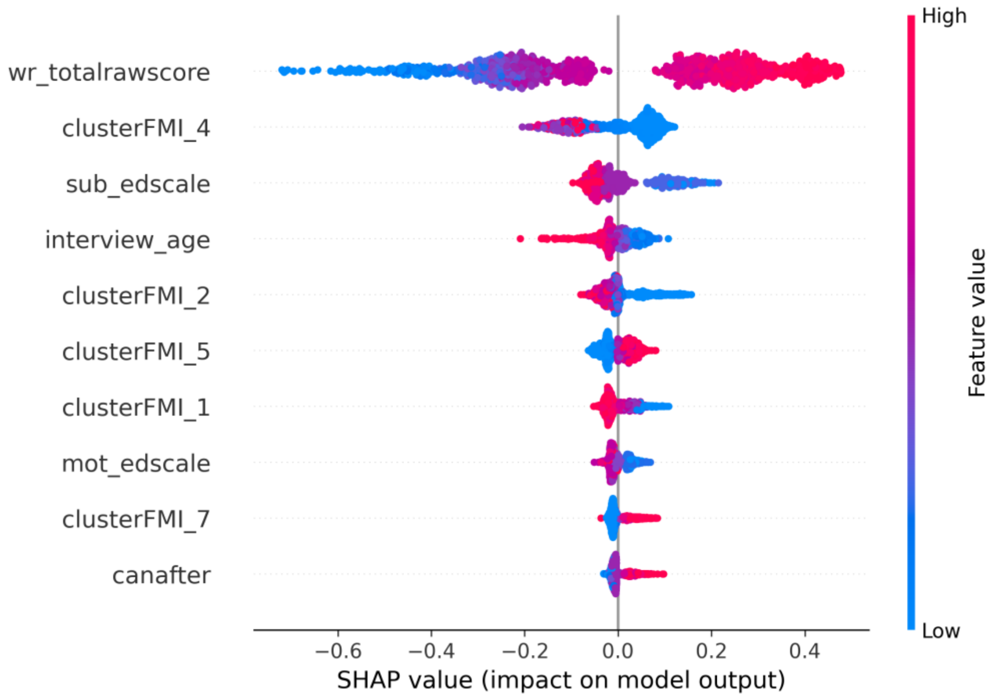


**S8A. SHAP summary plot**

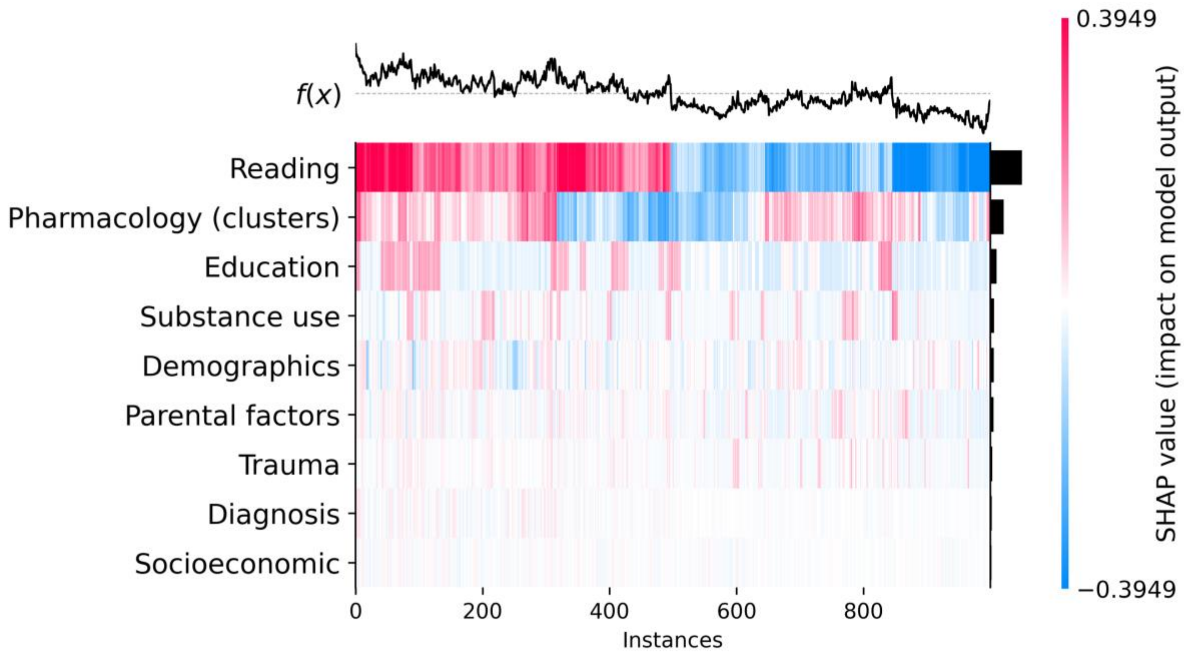


**S8B: Grouped SHAP heatmap**

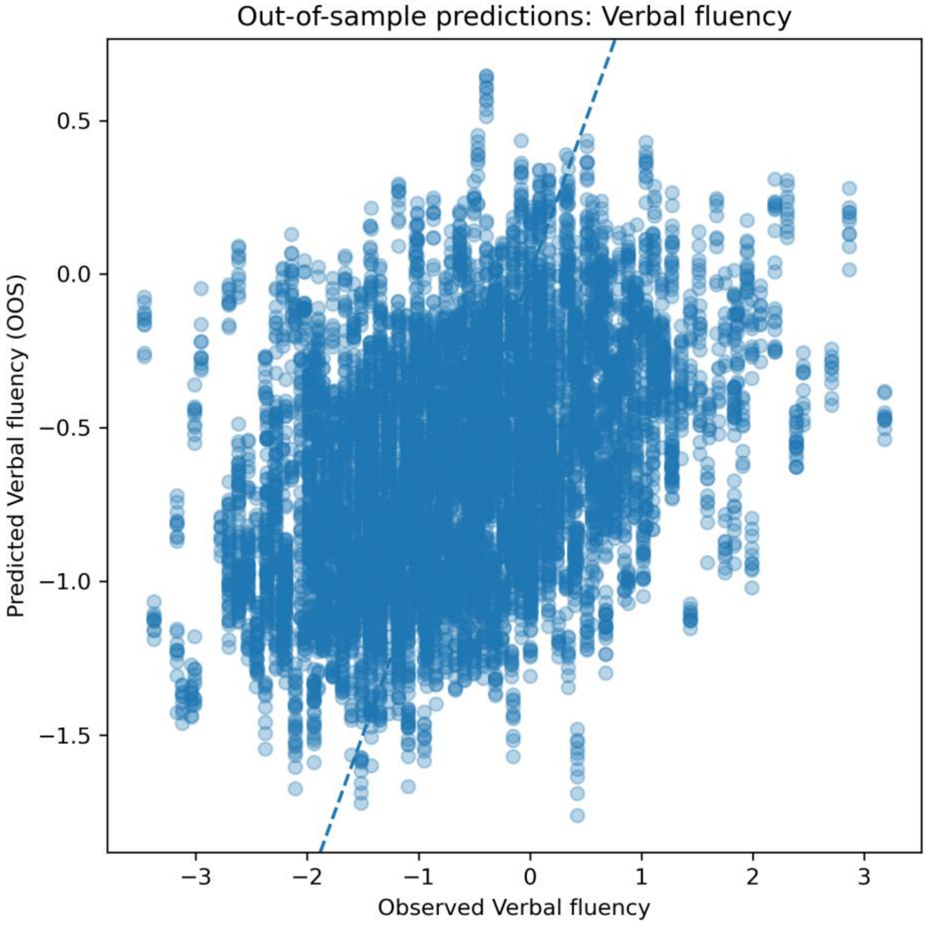


**S8C: Out-of-sample predicted versus observed verbal fluency**

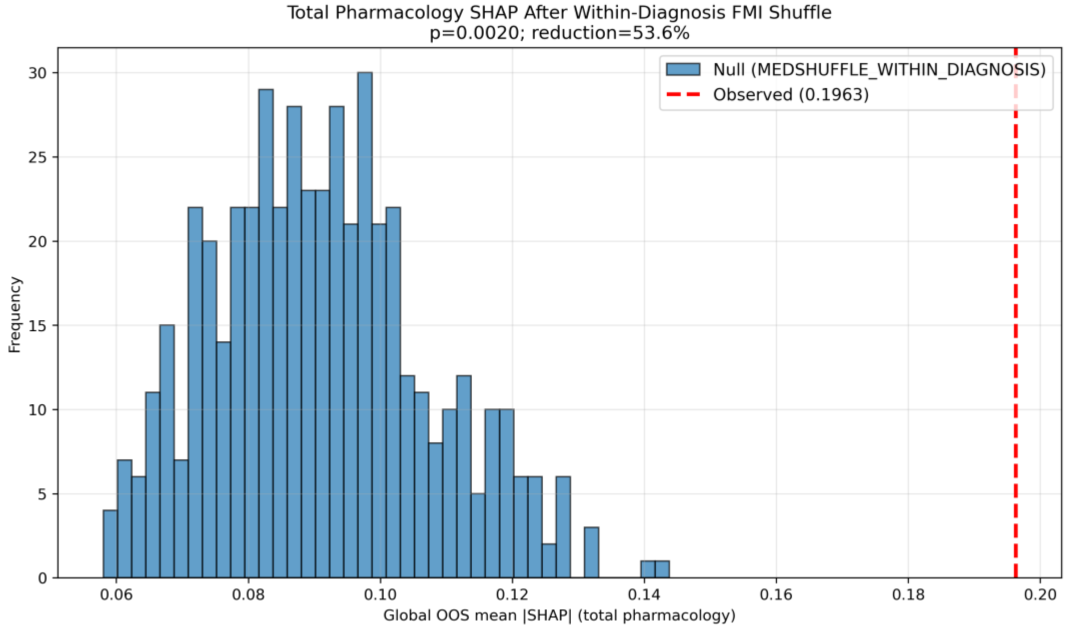


**S8D: Total pharmacology permutation**

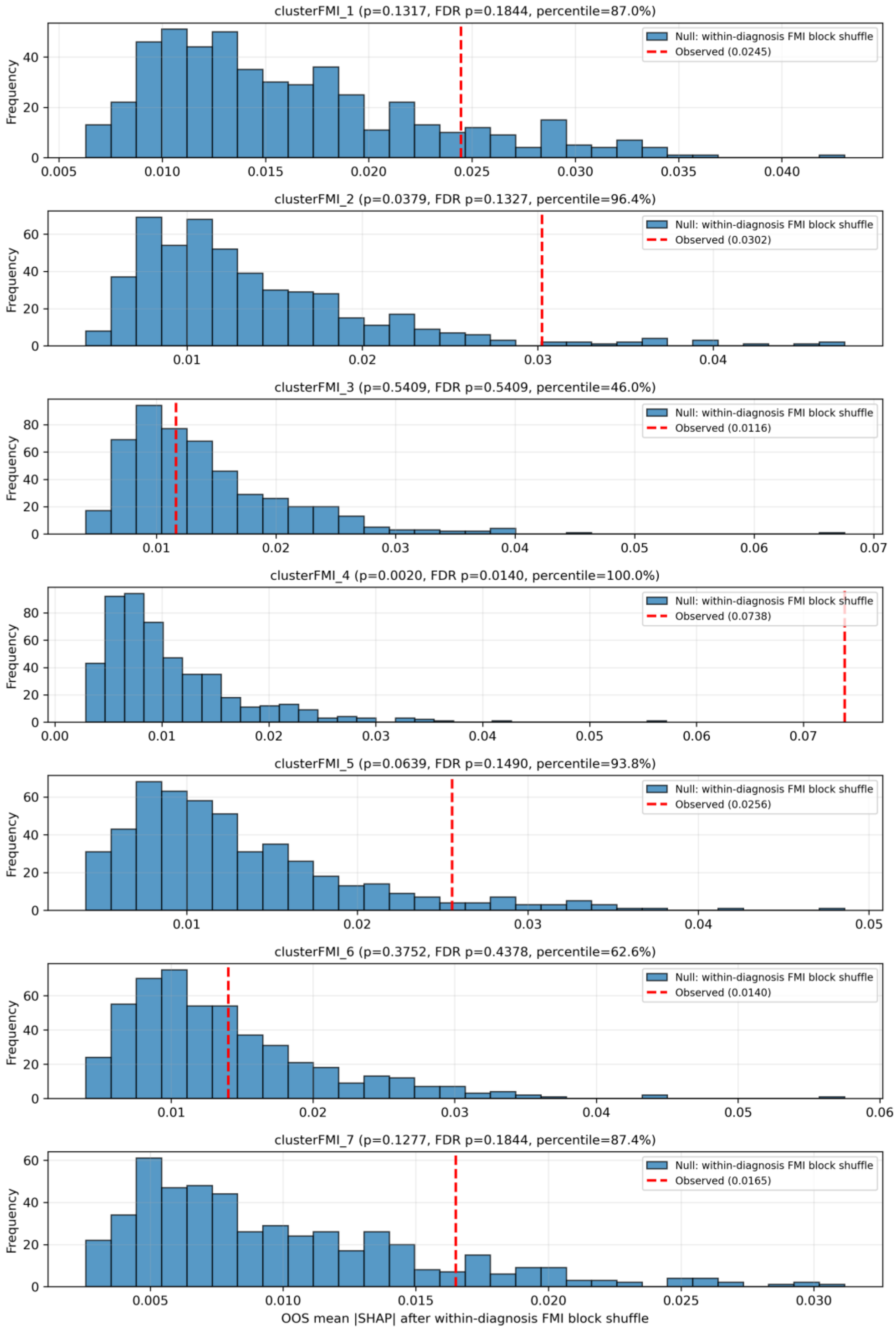


**S8E: Cluster-specific permutation null distributions**

**Supplementary Figure S9. Full covariate model - diagnostics and attribution on processing speed.**

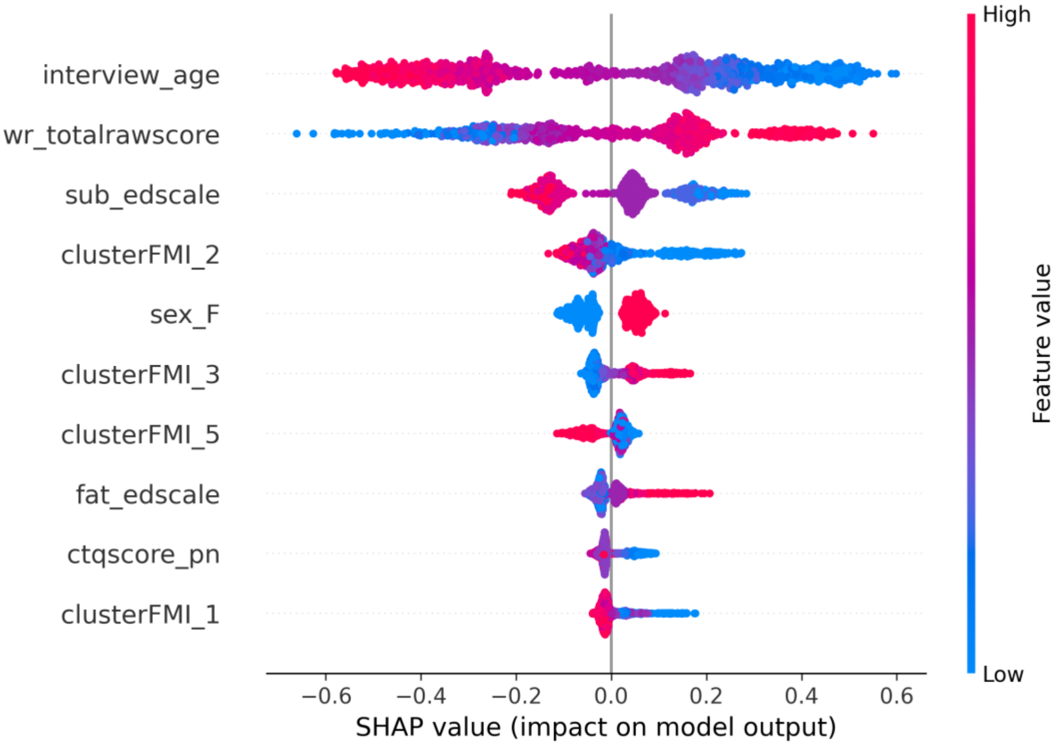


**S9A. SHAP summary plot**

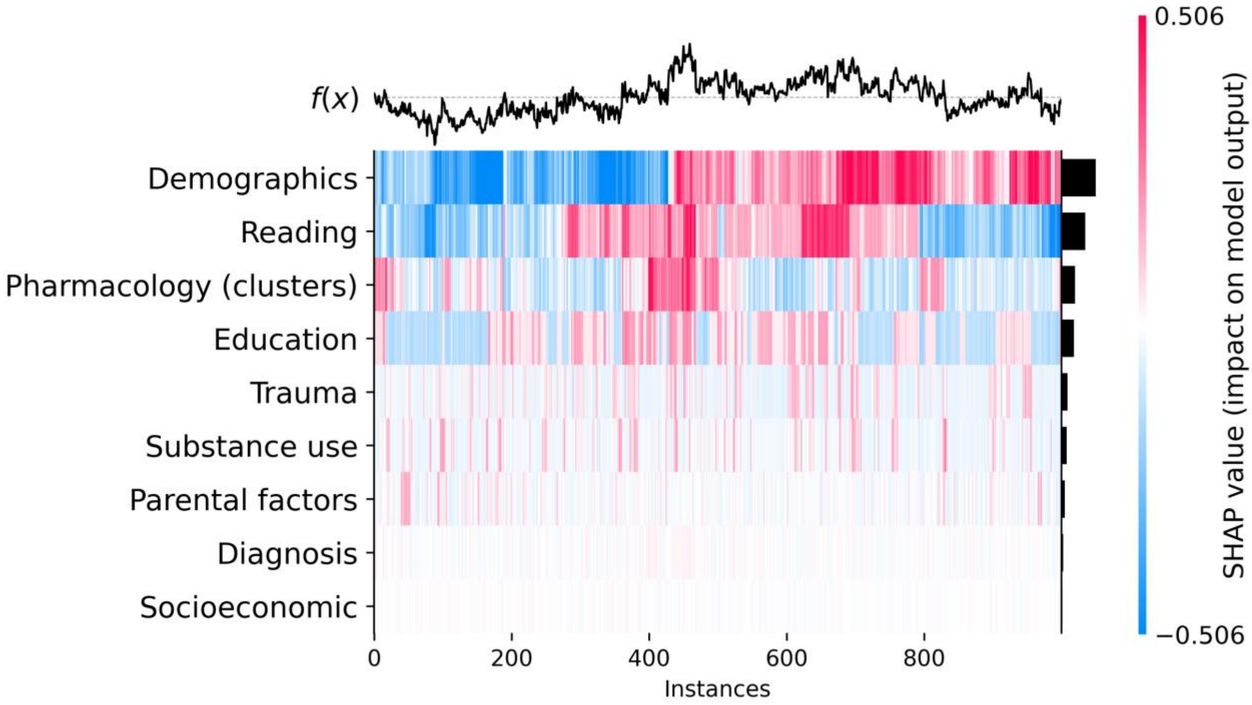


**S9B: Grouped SHAP heatmap**

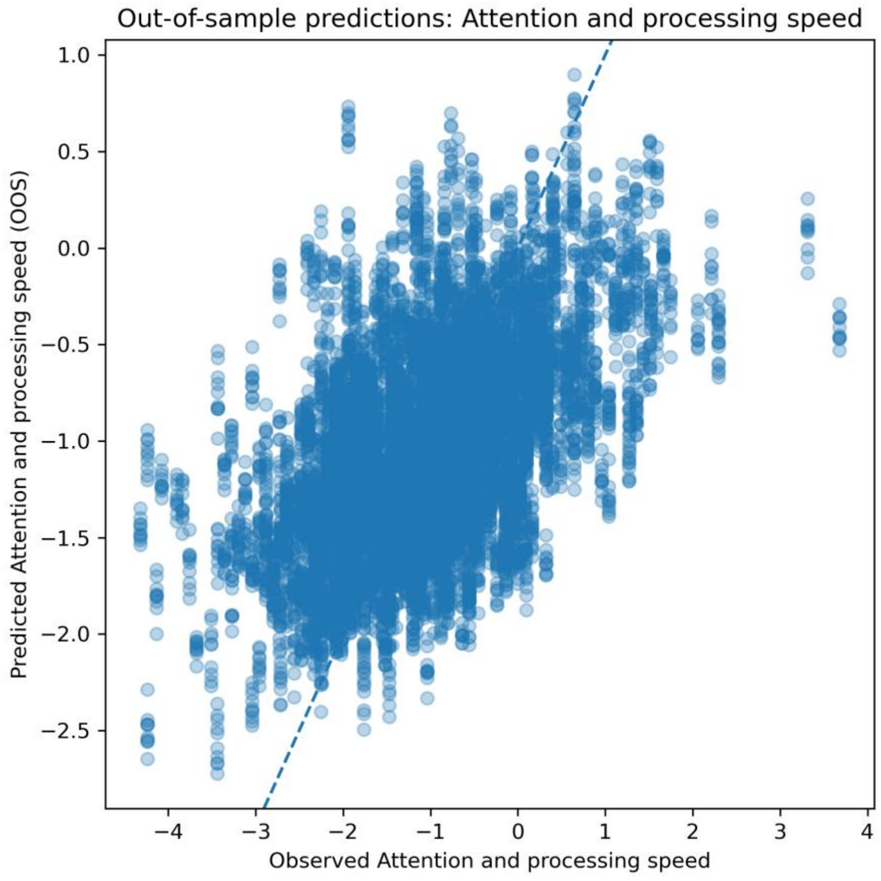


**S9C: Out-of-sample predicted versus observed processing speed**

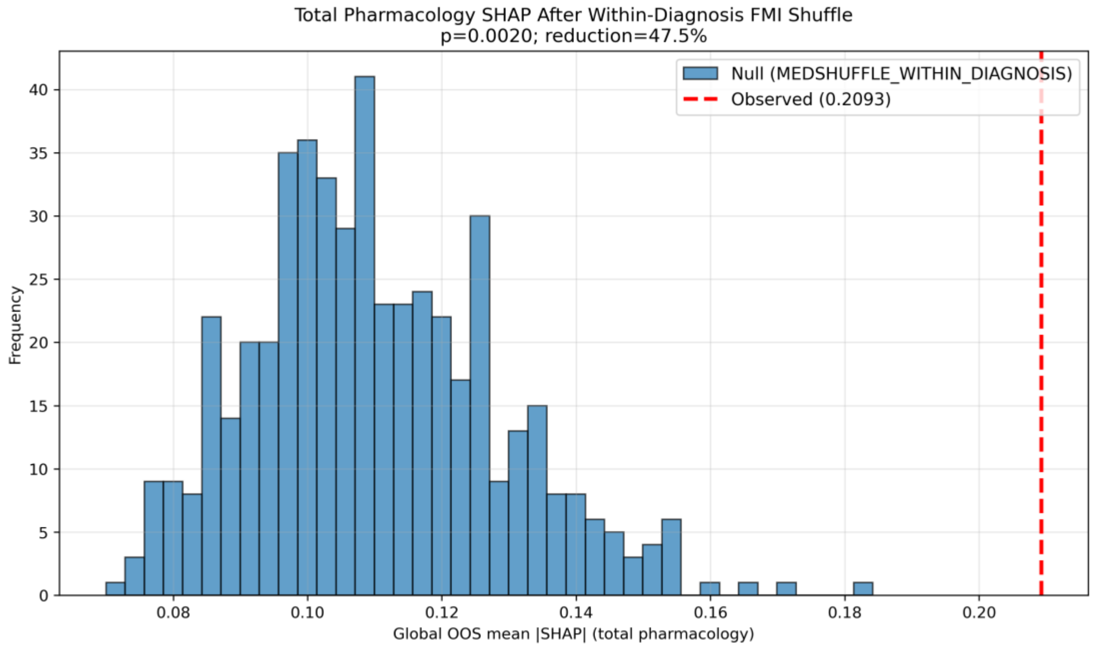


**S9D: Total pharmacology permutation**

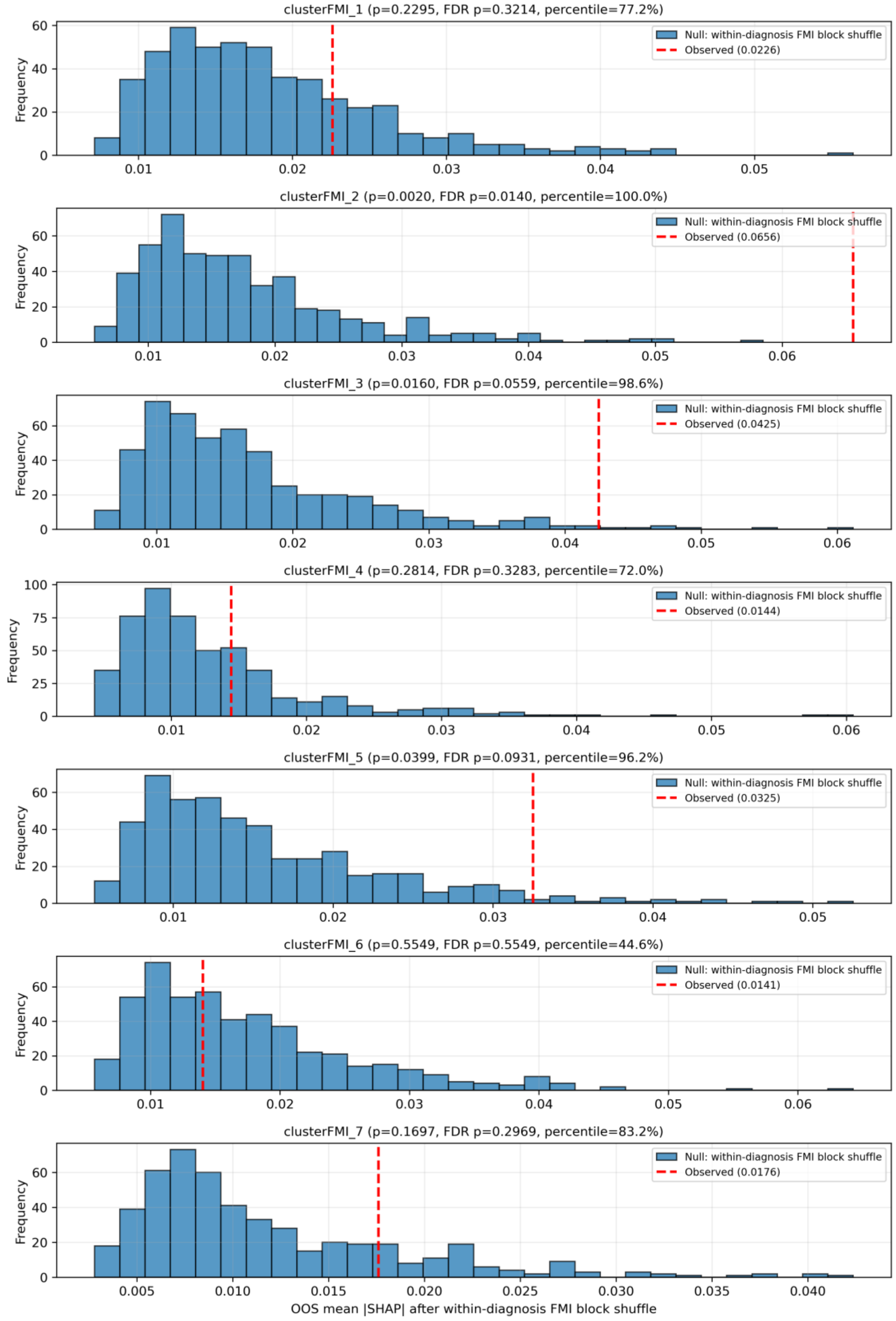


**S9E: Cluster-specific permutation null distributions**

**Supplementary Figure S10. Full covariate model - diagnostics and attribution on executive function.**

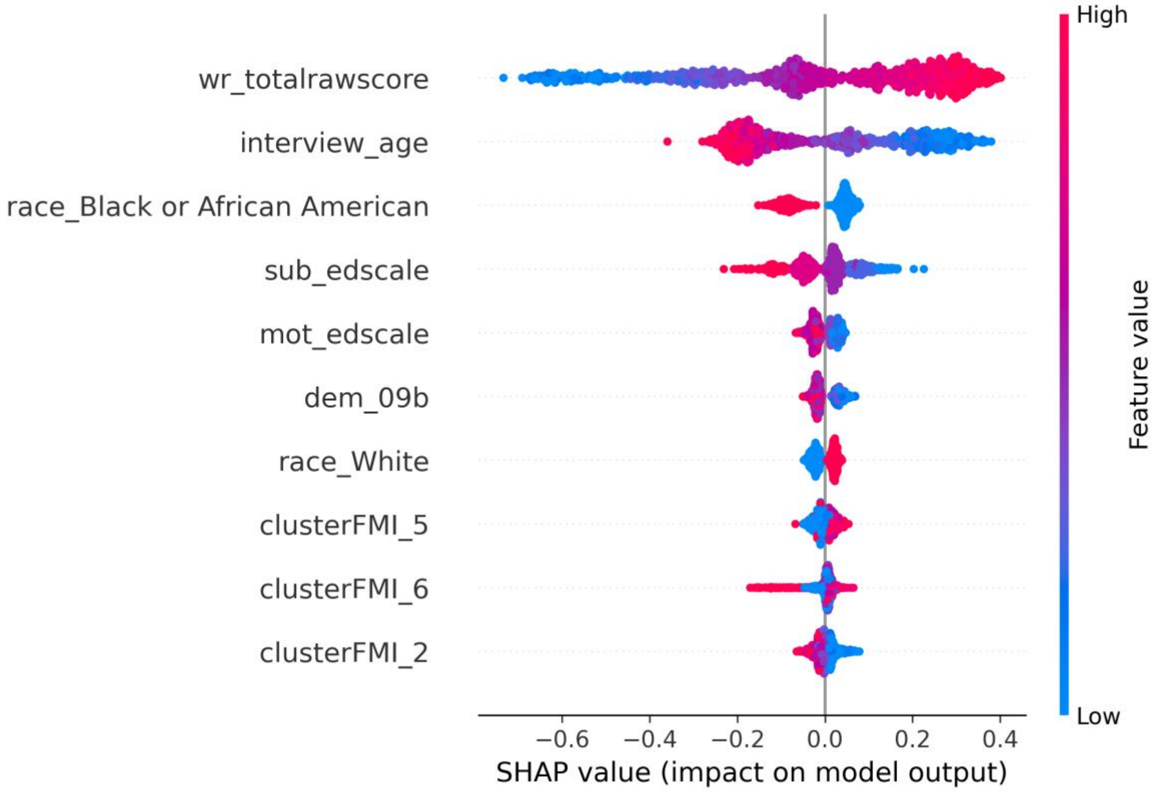


**S10A. SHAP summary plot**

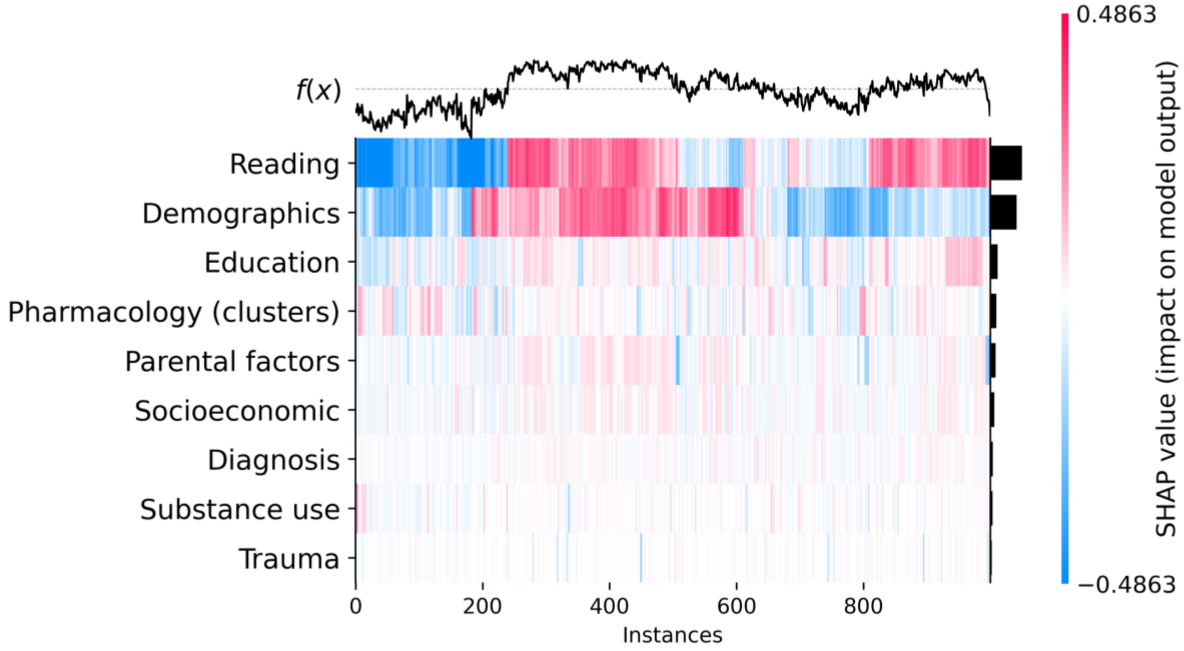


**S10B: Grouped SHAP heatmap**

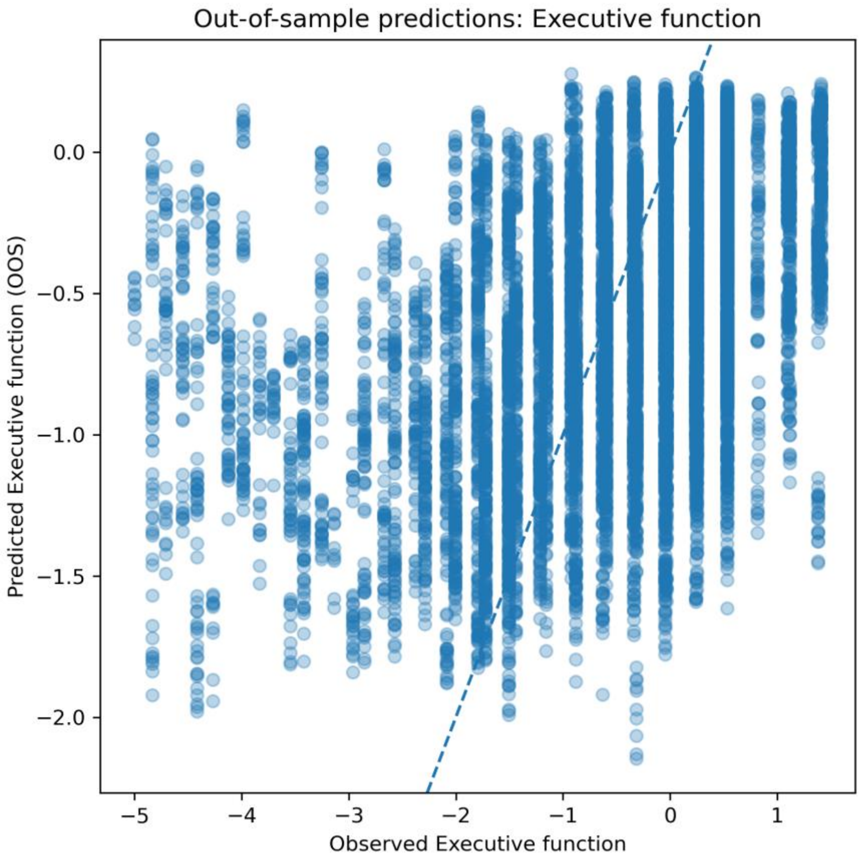


**S10C: Out-of-sample predicted versus observed executive function**

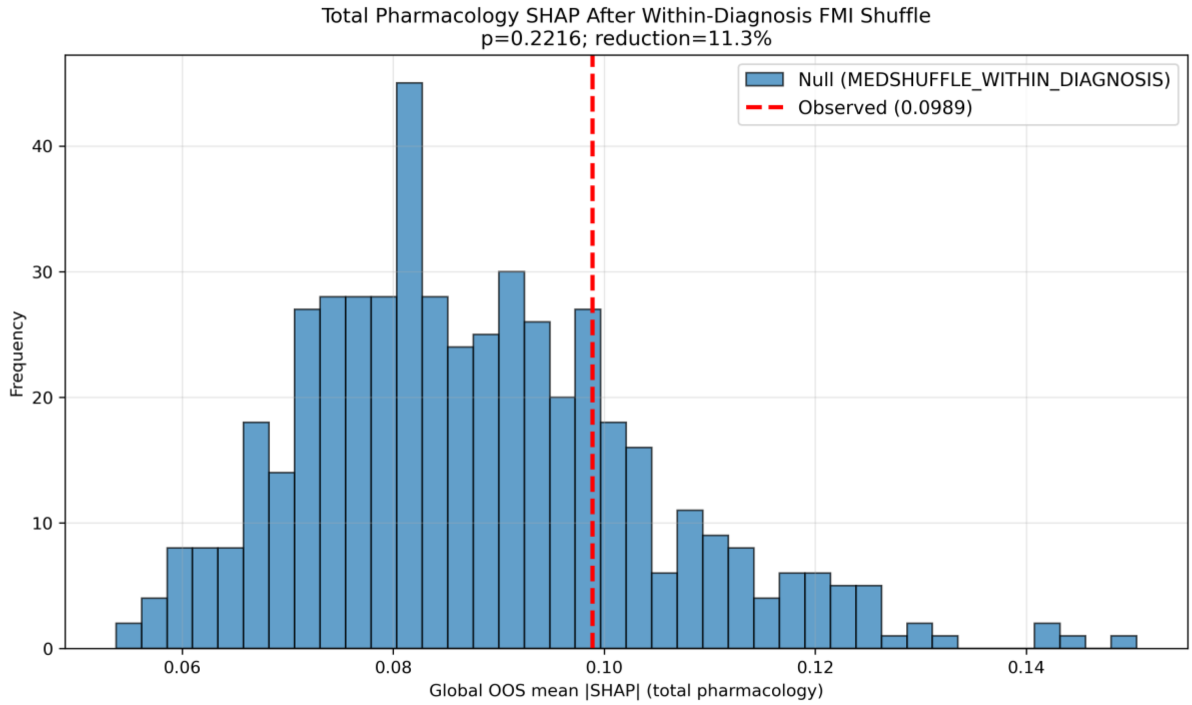


**S10D: Total pharmacology permutation**

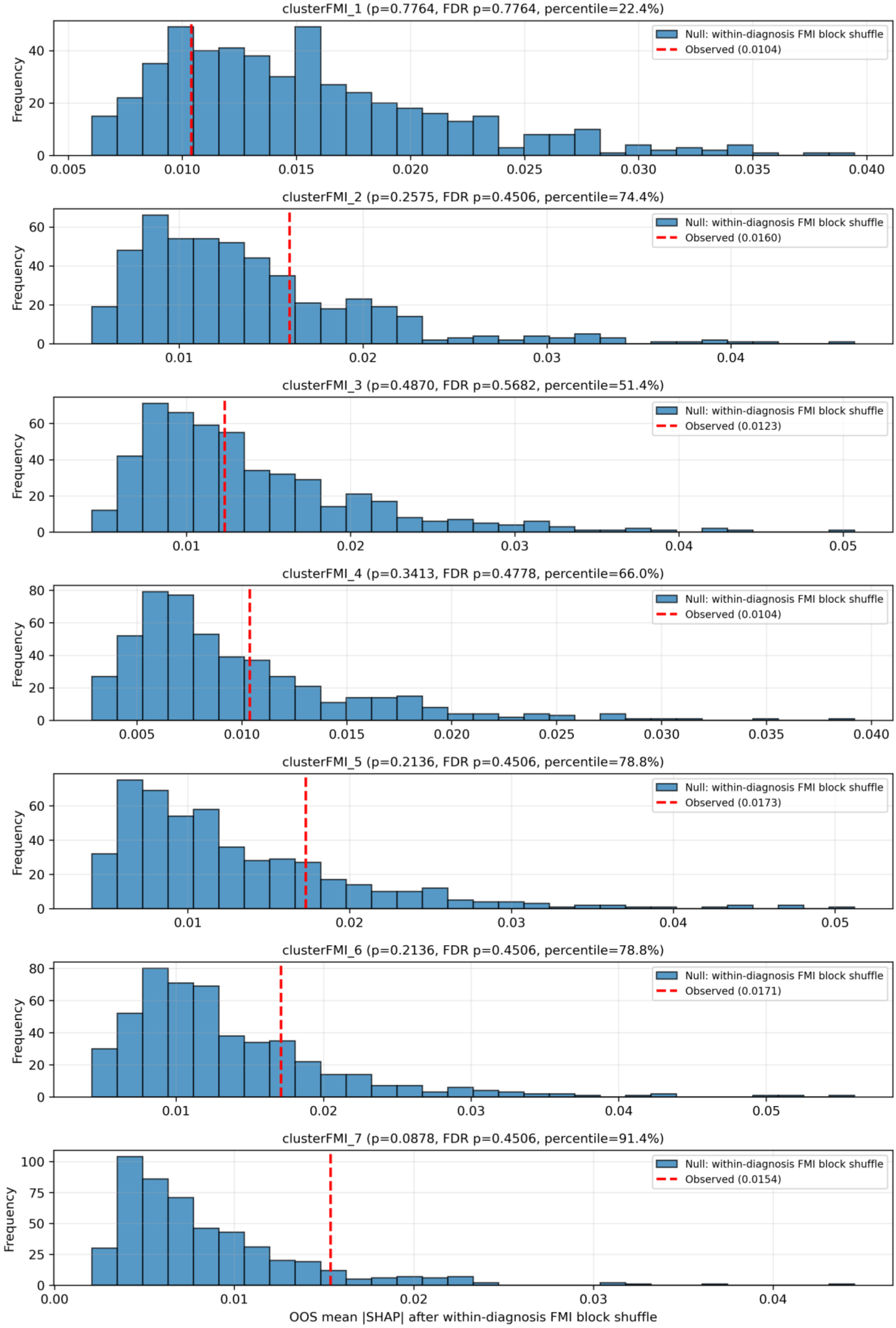


**S10E: Cluster-specific permutation null distributions**
